# Novel genomic loci influence patterns of structural covariance in the human brain

**DOI:** 10.1101/2022.07.20.22277727

**Authors:** Junhao Wen, Ilya M. Nasrallah, Ahmed Abdulkadir, Theodore D. Satterthwaite, Zhijian Yang, Guray Erus, Timothy Robert-Fitzgerald, Ashish Singh, Aristeidis Sotiras, Aleix Boquet-Pujadas, Elizabeth Mamourian, Jimit Doshi, Yuhan Cui, Dhivya Srinivasan, Ioanna Skampardoni, Jiong Chen, Gyujoon Hwang, Mark Bergman, Jingxuan Bao, Yogasudha Veturi, Zhen Zhou, Shu Yang, Paola Dazzan, Rene S. Kahn, Hugo G. Schnack, Marcus V. Zanetti, Eva Meisenzahl, Geraldo F. Busatto, Benedicto Crespo-Facorro, Christos Pantelis, Stephen J. Wood, Chuanjun Zhuo, Russell T. Shinohara, Ruben C. Gur, Raquel E. Gur, Nikolaos Koutsouleris, Daniel H. Wolf, Andrew J. Saykin, Marylyn D. Ritchie, Li Shen, Paul M. Thompson, Olivier Colliot, Katharina Wittfeld, Hans J. Grabe, Duygu Tosun, Murat Bilgel, Yang An, Daniel S. Marcus, Pamela LaMontagne, Susan R. Heckbert, Thomas R. Austin, Lenore J. Launer, Mark Espeland, Colin L Masters, Paul Maruff, Jurgen Fripp, Sterling C. Johnson, John C. Morris, Marilyn S. Albert, R. Nick Bryan, Susan M. Resnick, Yong Fan, Mohamad Habes, David Wolk, Haochang Shou, Christos Davatzikos, the iSTAGING, the BLSA, the BIOCARD, the PHENOM, the ADNI studies, the AI4AD consortium

## Abstract

Normal and pathologic neurobiological processes influence brain morphology in coordinated ways that give rise to patterns of structural covariance (PSC) across brain regions and individuals during brain aging and diseases. The genetic underpinnings of these patterns remain largely unknown. We apply a stochastic multivariate factorization method to a diverse population of 50,699 individuals (12 studies, 130 sites) and derive data-driven, multi-scale PSCs of regional brain size. PSCs were significantly correlated with 915 genomic loci in the discovery set, 617 of which are novel, and 72% were independently replicated. Key pathways influencing PSCs involve reelin signaling, apoptosis, neurogenesis, and appendage development, while pathways of breast cancer indicate potential interplays between brain metastasis and PSCs associated with neurodegeneration and dementia. Using support vector machines, multi-scale PSCs effectively derive imaging signatures of several brain diseases. Our results elucidate new genetic and biological underpinnings that influence structural covariance patterns in the human brain.

**Significance statement:** The coordinated patterns of changes in the human brain throughout life, driven by brain development, aging, and diseases, remain largely unexplored regarding their underlying genetic determinants. This study delineates 2003 multi-scale patterns of structural covariance (PSCs) and identifies 617 novel genomic loci, with the mapped genes enriched in biological pathways implicated in reelin signaling, apoptosis, neurogenesis, and appendage development. Overall, the 2003 PSCs provide new genetic insights into understanding human brain morphological changes and demonstrate great potential in predicting various neurologic conditions.

## Introduction

Brain structure and function are interrelated via complex networks that operate at multiple scales, ranging from cellular and synaptic processes, such as neural migration, synapse formation, and axon development, to local and broadly connected circuits.^1^ Due to a fundamental relationship between activity and structure, many normal and pathologic neurobiological processes, driven by genetic and environmental factors, collectively cause coordinated changes in brain morphology. Structural covariance analyses investigate such coordinated changes by seeking patterns of structural covariation (PSC) across brain regions and individuals.^1^ For example, during adolescence, PSCs derived from magnetic resonance imaging (MRI) have been considered to reflect a coordinated cortical remodeling as the brain establishes mature networks of functional specialization.^2^ Structural covariance is not only related to normal brain development or aging processes but can also reflect coordinated brain change due to disease. For example, individuals with motor speech dysfunction may develop brain atrophy in Broca’s inferior frontal cortex and co-occurring brain atrophy in Wernicke’s area of the superior temporal cortex.^3^ Refer to **Fig. 1C** for an illustrative depiction.

**Figure 1:**
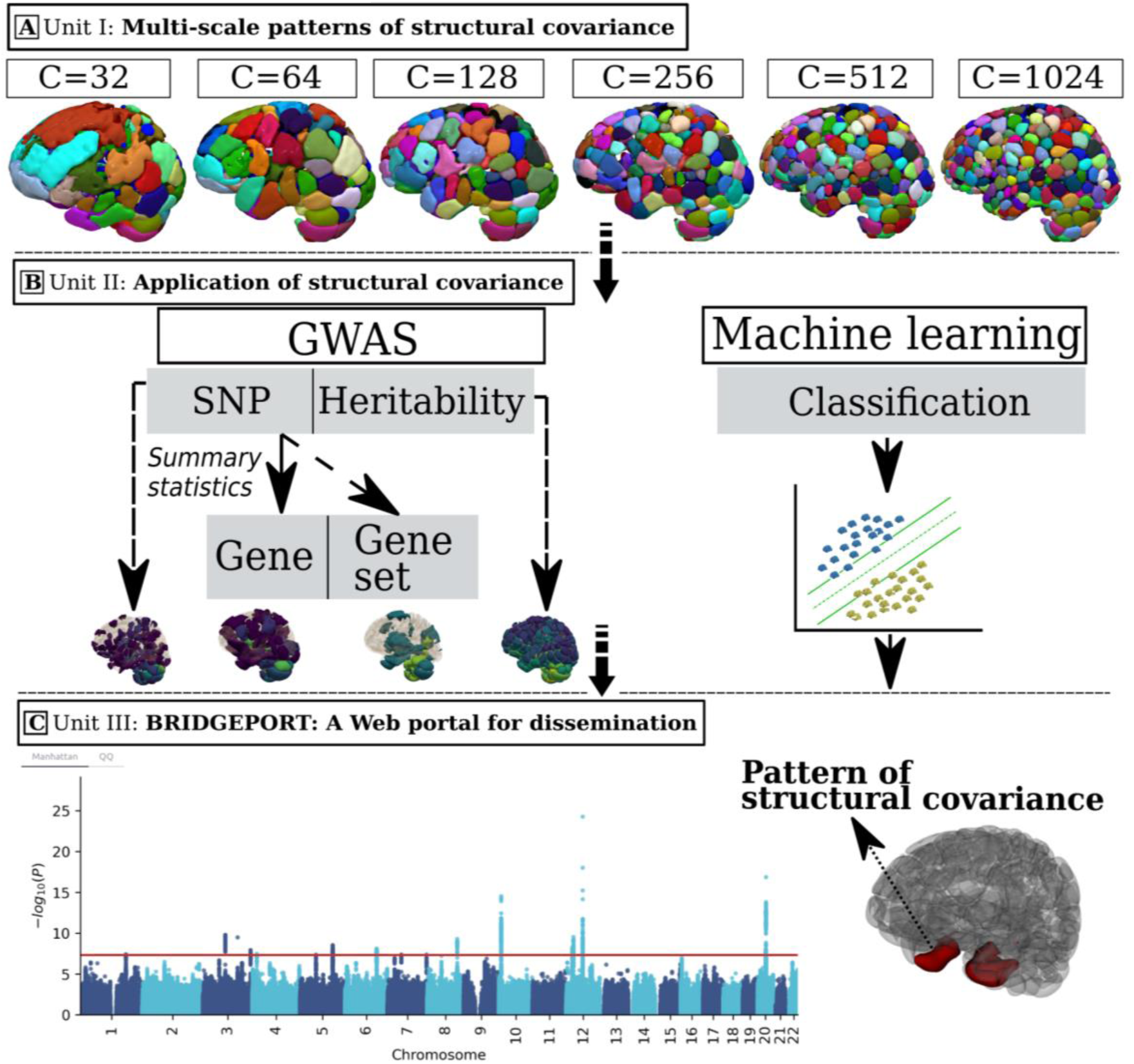
Study workflow. **A)** Unit I: the stochastic orthogonally projective non-negative matrix factorization (sopNMF) algorithm was applied to a large, disease-diverse population to derive multi-scale patterns of structural covariance (PSC) at different scales (*C*=32, 64, 128, 256, 512, and 1024; *C* represents the number of PSCs). **B**) Unit II: two types of analyses were performed in this study: Genome-wide association studies (GWAS) relate each of the PSCs (*N*=2003) to common genetic variants; pattern analysis via machine learning demonstrates the utility of the multi-scale PSCs in deriving individualized imaging signatures of various brain pathologies. **C**) Unit III: BRIDGEPORT is a web portal that makes all resources publicly available for dissemination. As an illustration, a Manhattan plot for PSC (C64-3, the third PSC of the C64 atlas) and its 3D brain map are displayed.

The human brain develops, matures, and degenerates in coordinated patterns of structural covariance at the macrostructural level of brain morphology.^1^ However, the mechanisms underlying structural covariance are still unclear, and their genetic underpinnings are largely unknown. We hypothesized that brain morphology was driven by multiple genes (i.e., polygenic) collectively operating on different brain areas (i.e., pleiotropic), resulting in connected networks covaried by normal aging and various disease-related processes. Along the causal pathway from underlying genetics to brain morphological changes, we sought to elucidate which genetic underpinnings (e.g., genes), biological processes (e.g., neurogenesis), cellular components (e.g., nuclear membrane), molecular functions (e.g., nucleic acid binding), and neuropathological processes (e.g., Alzheimer’s disease) might influence the formation, development, and changes of structural covariance patterns in the human brain.

Previous neuroimaging genome-wide association studies (GWAS)^4, 5^ have partially investigated the abovementioned questions and expanded our understanding of the genetic architecture of the human brain. However, they focused on conventional neuroanatomical regions of interest (ROI) instead of data-driven PSCs. In brain imaging research, prior studies have applied structural covariance analysis to elucidate underlying coordinated morphological changes in brain aging and various brain diseases,^1^ but have had several limitations. They often relied on pre-defined neuroanatomical ROIs to construct inter- and intra-individual structural covariance networks. These *a priori* ROIs might not optimally reflect the molecular-functional characteristics of the brain. In addition, most population-based studies have investigated brain structural covariance within a relatively limited scope, such as within relatively small samples, over a relatively narrow age window (e.g., adolescence^2^), within a single disease (e.g., Parkinson’s disease^6^), or within datasets lacking sufficient diversity in cohort characteristics or MRI scanner protocols. These have been imposed, in part, by limitations in both available cohort size and in the algorithmic implementation of structural covariance analysis, which has been computationally restricted to modest sample sizes when investigated at full image resolution. Lastly, prior studies have examined brain structural covariance at a single fixed ROI resolution/scale/granularity. While the optimal scale is unknown and may differ by the question of interest, the highly complex organization of the human brain may demonstrate structural covariance patterns that span multiple scales.^7, 8^

To address this gap, we modified our previously proposed orthogonally projective non-negative matrix factorization (opNMF^9^) to its stochastic counterpart, sopNMF. This adaptation allowed us to train the model iteratively on large-scale neuroimaging datasets with a pre-defined number of PSCs (*C*). Non-negative matrix factorization has gained significant attention in neuroimaging due to its ability to reduce complex data into a sparse, part-based brain representation by projection onto a relatively small number of components (the PSCs). NMF has been shown to substantially improve interpretability and reproducibility compared to other unsupervised methods, such as PCA and ICA, thanks to the non-negative constraint that produces parcellation-like decompositions of complex signals. Our opNMF/sopNMF approach imposed an additional orthonormality constraint^9^ (*Equation 1* in **Method 1**), further enhancing sparsity and facilitating clinical interpretability. In our previous work, we applied the opNMF method to 934 youths ages 8–20 to depict the coordinated growth of structural brain networks during adolescence – a period characterized by extensive remodeling of the human cortex to accommodate the rapid expansion of the behavioral repertoire^2^. Remarkably, this study revealed PSCs that exhibited a cortical organization closely aligned with established functional brain networks, such as the well-known 7-network functional parcellation proposed by Yeo et al^10^. Notably, this alignment emerged without prior assumptions, was data-driven and hypothesis-free, and potentially reflected underlying neurobiological processes related to brain development and aging. Herein, we used large-scale neuroimaging data to investigate the underlying genetic determinant influencing such changes in structural covariance patterns in the human brain.

We examined structural covariance of regional cortical and subcortical volume in the human brain using MRI from a diverse population of 50,699 people from 12 studies, 130 sites, and 12 countries, comprised of cognitively healthy individuals, as well as participants with various diseases/conditions over their lifespan (ages 5 through 97). Herein we present results from coarse to fine scales corresponding to *C* = 32, 64, 128, 256, 512, and 1024. We hypothesized that PSCs at multiple scales could delineate the human brain’s multi-factorial and multi-faceted morphological landscape and genetic architecture in healthy and diseased individuals. We examined the associations between these multi-scale PSCs and common genetic variants at different levels (*N*=8,469,833 SNPs). In total, 617 novel genomic loci were identified; key pathways (e.g., neurogenesis and reelin signaling) contributed to shaping structural covariance patterns in the human brain. In addition, we leveraged PSCs at multiple scales to better derive individualized imaging signatures of several diseases than any single-scale PSCs using support vector machines. All experimental results and the multi-scale PSCs were integrated into the MuSIC (Multi-scale Structural Imaging Covariance) atlas and made publicly accessible through the BRIDGEPORT (**BR**aIn know**l**e**DGE PORT**al) web portal: https://www.cbica.upenn.edu/bridgeport/. **Table 1** provides an overview of the abbreviations used in the present study.

**Table 1.** Abbreviations used in the present study.

| Item | Abbreviation | Item | Abbreviation |
| --- | --- | --- | --- |
| Pattern of structural covariation | PSC | Independent component analysis | ICA |
| Genome-wide association study | GWAS | BRaIn knowleDGE PORTal | BRIDGEPORT |
| Orthogonal projective non-negative matrix factorization | opNMF | Multi-scale Structural Imaging Covariance | MuSIC |
| Stochastic orthogonal projective non-negative matrix factorization | sopNMF | Machine learning | ML |
| Principal component analysis | PCA | UK Biobank | UKBB |
| Imaging-based coordinate SysTem for AGing and NeurodeGenerative diseases | iSTAGING | Psychosis Heterogeneity Evaluated via Dimensional Neuroimaging | PHENOM |
| Single nucleotide polymorphism | SNP | Region of interest | ROI |
| Magnetic resonance imaging | MRI | Automated anatomical labeling | AAL |
| MULTi-atlas region Segmentation utilizing Ensembles | MUSE | Alzheimer's disease | AD |
| Spatial PATterns for REcognition | SPARE | Support vector machine | SVM |

## Results

We summarize this work in three units (I to III) outlined in **Fig. 1**. In Unit **I** (**Fig. 1A**), we present the stochastic orthogonally projective non-negative matrix factorization (sopNMF) algorithm (**Method 1**), optimized for large-scale multivariate structural covariance analysis. The sopNMF algorithm decomposes large-scale imaging data through online learning to overcome the memory limitations of opNMF. A subgroup of participants with multiple disease diagnoses and healthy controls (ages 5-97, training population, *N*=4000, **Method 2**) were sampled from the discovery set (*N*=32,440, **Method 2**); their MRI underwent a standard imaging processing pipeline (**Method 3A**). The processed images were then fit to sopNMF to derive the multi-scale PSCs (*N*=2003) from the loadings of the factorization (**Method 1**). We incorporate participants with various disease conditions because previous studies have demonstrated that inter-regional correlated patterns (i.e., depicting a network) show variations in healthy and diseased populations, albeit to a differing degree.^11^ Multi-scale PSCs were extracted across the entire population and statistically harmonized^12^ (**Method 3B**). Unit **II** (**Fig. 1B**) investigates the harmonized data for 2003 PSCs (13 PSCs have vanished in this process for *C*=1024; see **Method 1**) in two brain structural covariance analyses. Specifically, we performed *i*) GWAS (**Method 4**) that sought to discover associations of PSCs at single nucleotide polymorphism (SNP), gene, or gene set-level; and *ii*) pattern analysis via support vector machine (**Method 5**) to derive individualized imaging signatures of several brain diseases and conditions. Unit **III** (**Fig. 1C**) presents BRIDGEPORT, making these massive analytic resources publicly available to the imaging, genomics, and machine learning communities.

### Patterns of structural covariance via stochastic orthogonally projective non-negative matrix factorization

We first validated the sopNMF algorithm by showing that it converged to the global minimum of the factorization problem using the comparison population (*N*=800, **Method 2**). The sopNMF algorithm achieved similar reconstruction loss and sparsity as opNMF but at reduced memory demand (**Supplementary eFigure 1**). The lower memory requirements of sopNMF made it possible to generate multi-scale PSCs by jointly factorizing 4000 MRIs in the training population. The results of the algorithm were robust and obtained a high reproducibility index (RI) (**Supplementary eMethod 2**) in several reproducibility analyses: split-sample analysis (RI = 0.76±0.27), split-sex analysis (RI = 0.79±0.27), and leave-one-site-out analysis (RI = 0.65-0.78 for C32 PSCs) (**Supplementary eFigure 2**). We then extracted the multi-scale PSCs in the discovery set (*N*=32,440) and the replication set (*N*=18,259, **Method 2**) for Unit II. These PSCs succinctly capture underlying neurobiological processes across the lifespan, including the effects of typical aging processes and various brain diseases. In addition, the multi-scale representation constructs a hierarchy of brain structure networks (e.g., PSCs in cerebellum regions), which models the human brain in a multi-scale topology.^7, 13^

### Patterns of structural covariance are highly heritable

The multi-scale PSCs are highly heritable (0.05< *h*^2^ *<*0.78), showing high SNP-based heritability estimates (*h*^2^) (**Method 4B**) for the discovery set (**Fig. 2**). Specifically, the *h*^2^ estimate was 0.49±0.10, 0.39±0.14, 0.29±0.15, 0.25±0.15, 0.27±0.15, 0.31±0.15 for scales *C*=32, 64, 128, 256, 512 and 1024 of the PSCs, respectively. The Pearson correlation coefficient between the two independent estimates of *h*^2^ was *r* = 0.94 (p-value < 10^-6^, between the discovery and replication sets) in the UK Biobank (UKBB) data. The scatter plot of the two sets of *h^2^* estimates is shown in **Supplementary eFigure 3**. The *h^2^* estimates and p-values for all PSCs are detailed in **Supplementary eFile 1** (discovery set) and **eFile 2** (replication set). Our results confirm that brain structure is heritable to a large extent and identify the spatial distribution of the most highly heritable regions of the brain (e.g., subcortical gray matter structures and cerebellum regions).^14^

**Figure 2:**
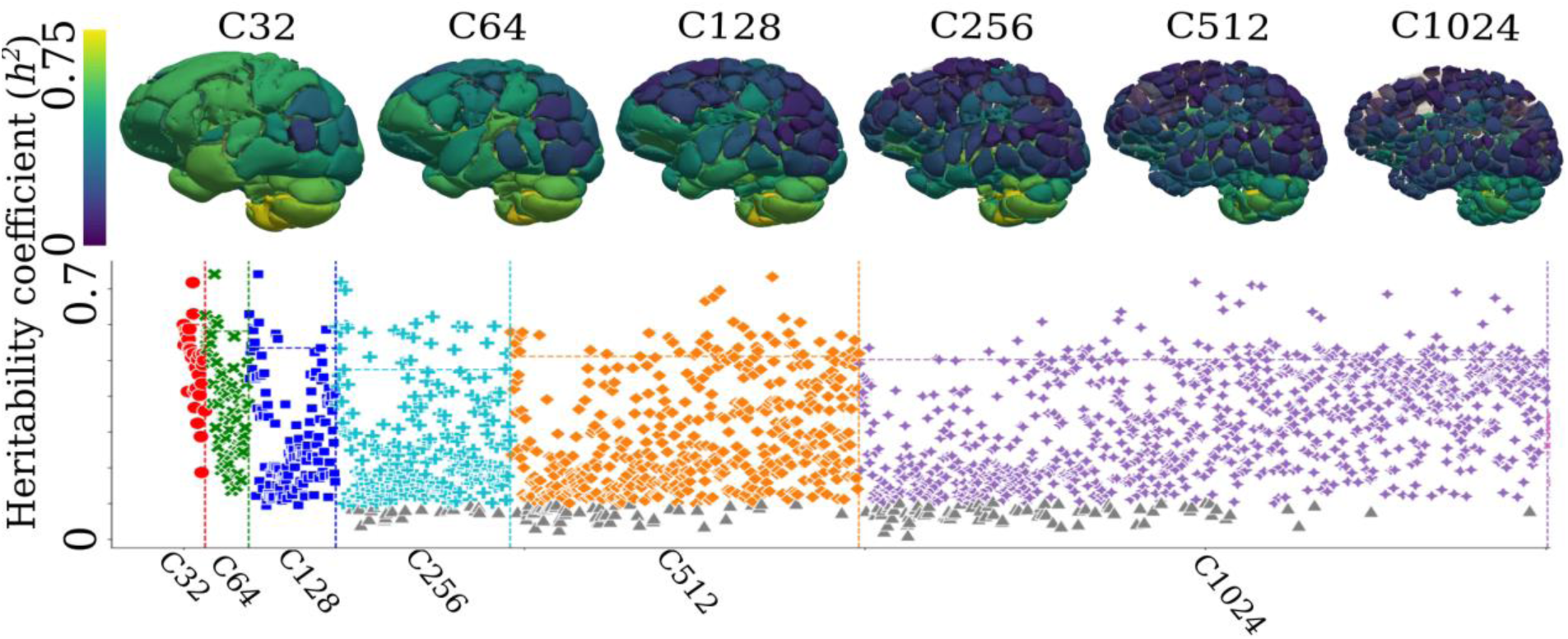
Patterns of structural covariance are highly heritable in the human brain. Patterns of structural covariance (PSCs) of the human brain are highly heritable. The SNP-based heritability estimates are calculated for the multi-scale PSCs at different scales (*C*). PSCs surviving Bonferroni correction for multiple comparisons are depicted in color in the Manhattan plots (gray otherwise). Each PSC’s heritability estimate (h2) was projected onto the 3D image space to show a statistical map of the brain at each scale *C*. The dotted line indicates each scale’s top 10% of most heritable PSCs.

### 617 novel genomic loci of patterns of structural covariance

We discovered genomic locus-PSC pairwise associations (**Method 4C**, **Supplementary eMethod 5**) within the discovery set and then independently replicated these associations on the replication set. We found that 915 genomic loci had 3791 loci-PSC pairwise significant associations with 924 PSCs after Bonferroni correction (**Method 4G**) for the number of PSCs (p-value threshold per scale: 10.3 > -log_10_[p-value] > 8.8) (**Supplementary eFile 3**, and **Fig. 3A**). Our results showed that the formation of these PSCs is largely polygenic; the associated SNPs might play a pleiotropic role in shaping these networks.

**Figure 3:**
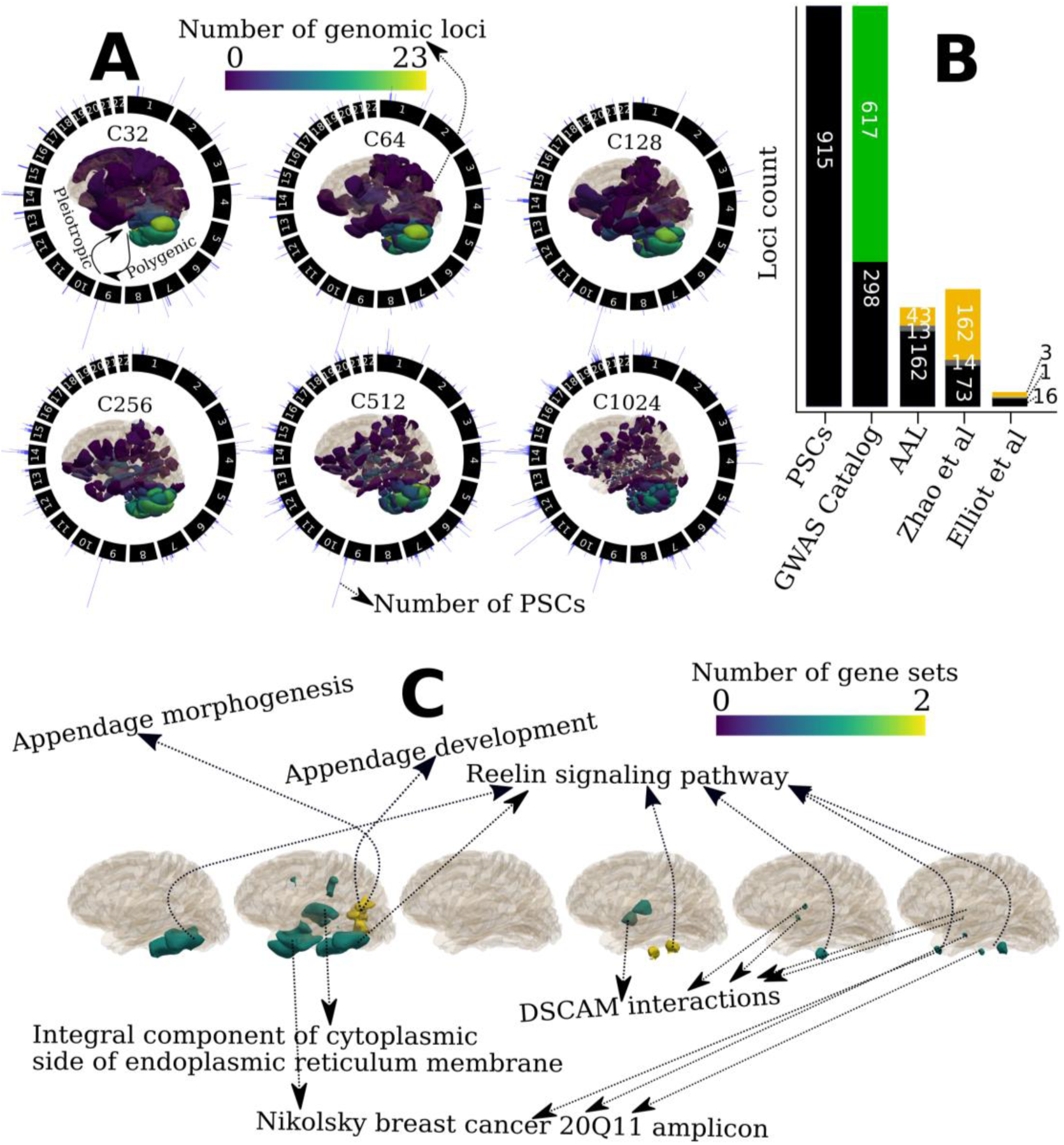
Patterns of structural covariance highlight novel genomic loci and pathways that shape the human brain. **A**) Patterns of structural covariance (PSC) in the human brain are polygenic: the number of genomic loci of each PSC is projected onto the image space to show a statistical brain map characterized by the number (*C*) of PSCs. In addition, common genetic variants exert pleiotropic effects on the PSCs: circular plots showed the number of associated PSCs (histograms in blue color) of each genomic loci over the entire autosomal chromosome (1-22). The histogram was plotted for the number of PSCs for each genomic locus in the circular plots. **B**) Novel genomic loci revealed by the multi-scale PSCs compared to previous findings from the GWAS Catalog,^15^ T1-weighted MRI GWAS^4,5^, and the AAL atlas regions of interest. The green bar indicates the 617 novel genomic loci not previously associated with any clinical traits in GWAS Catalog; the black bar presents the loci identified in other studies that overlap (grey bar for loci in linkage disequilibrium) with the loci from our results; the yellow bar indicates the unique loci in other studies. **C**) Pathway enrichment analysis highlights six unique biological pathways and functional categories (after Bonferroni correction for 16,768 gene sets and the number of PSCs) that might influence the changes of PSCs. DSCAM: Down syndrome cell adhesion molecule.

Compared to previous literature, out of the 915 genomic loci, the multi-scale PSCs identified 617 novel genomic loci not previously associated with any traits or phenotypes in the GWAS Catalog^15^ (**Supplementary eFile 4**, **Fig. 3B**, query date: April 5^th^, 2023). These novel associations might indicate subtle neurobiological processes that are captured thanks to the biologically relevant structural covariance expressed by sopNMF. The multi-scale PSCs identified many novel associations by constraining this comparison to previous neuroimaging GWAS^12, 13^ using T1w MRI-derived phenotypes (e.g., regions of interest from conventional brain atlases) (**Fig 3B**, **Supplementary eTable 3**, **eFile 5**, **6**, and **7**).

Our UKBB replication set analysis (**Method 4H**) demonstrated that 3638 (96%) exact genomic locus-PSC associations were replicated at nominal significance (-log_10_[p-value] > 1.31), 2705 (72%) of which were significant after correction for multiple comparisons (**Method 4G,** - log_10_[p-value] > 4.27). We present this validation in **Supplementary eFile 8** from the replication set. The summary statistics, Manhattan, and QQ plots derived from the combined population (*N*=33,541) are presented in BRIDGEPORT.

In addition to the abovementioned replication analyses, we also performed several sensitivity analyses (**Supplementary e**Figure 4a). We used the GWAS results (233 significant SNPs in 5 genomic loci) of the first PSC in C32 (C32_1) from the UKBB discovery set to demonstrate this. First, we replicated all the 233 significant SNPs in 5 genomic loci both at the nominal level (-log_10_[p-value] > 1.31), and the Bonferroni corrected p-value threshold (-log_10_[p-value] > 3.67) using the combined discovery and replication sets (*N*=33,541) (**Supplementary eFigure 4b**), the 20,438 4ticipants with all ancestries in the discovery set (**Supplementary eFigure 4c**), and the 16,743 participants in the discovery set with four additional imaging-related covariates (3 parameters for the brain position in the lateral, longitudinal, and transverse directions, and 1 parameter for the head motion from fMRI) (**Supplementary eFigure 4d**). While replicating the results in 2386 participants with non-European ancestries, we only replicated 41 SNPs (17.6%), passing the nominal significant threshold (**Supplementary eFigure 4e**). Finally, only 14 SNPs (6.4%) were replicated when replicating the results using 1481 whole-genome sequencing (WGS) data from ADNI consolidated by the AI4AD consortium^16^ (**Supplementary eFigure 4f**). The low replication rates in other ancestries and independent disease-specific populations are expected due to population stratification, disease-specific effects, and reduced sample sizes. This further emphasizes the urge to enrich and diversify genetic research with non-European ancestries and disease-specific populations.

### Gene set enrichment analysis highlights pathways that shape patterns of structural covariance

For gene-level associations (**Method 4D**), we discovered that 164 genes had 2489 gene-PSC pairwise associations with 445 PSCs after Bonferroni correction for the number of genes and PSCs (p-value threshold: 8.6 > -log_10_[p-value] > 7.1) (**Supplementary eFile 9**).

Based on these gene-level p-values, we performed hypothesis-free gene set pathway analysis using MAGMA^17^(**Method 4E**): a more stringent correction for multiple comparisons was performed than the prioritized gene set enrichment analysis using *GENE2FUN* from FUMA (**Method 4F** and **Fig. 4**). We identified that six gene set pathways had 18 gene set-PSC pairwise associations with 17 PSCs after Bonferroni correction for the number of gene sets and PSCs (*N*=16,768 and *C* from 32 to 1024, p-value threshold: 8.54 > -log_10_[p-value] > 7.03) (**Fig. 3C**, **Supplementary eFile 10**). These gene sets imply critical biological and molecular pathways that might shape brain morphological changes and development. The reelin signaling pathway regulates neuronal migration, dendritic growth, branching, spine formation, synaptogenesis, and synaptic plasticity.^18^ The appendage morphogenesis and development pathways indicate how the anatomical structures of appendages are generated, organized, and progressed over time, often related to the cell adhesion pathway. These pathways elucidate how cells or tissues can be organized to create a complex structure like the human brain.^19^ In addition, the integral component of the cytoplasmic side of the endoplasmic reticulum membrane is thought to form a continuous network of tubules and cisternae extending throughout neuronal dendrites and axons.^20^ The DSCAM (Down syndrome cell adhesion molecule) pathway likely functions as a cell surface receptor mediating axon pathfinding. Related proteins are involved in hemophilic intercellular interactions.^21^ Lastly, Nikolsky et al.^22^ defined genes from the breast cancer 20Q11 amplicon pathway that were involved in the brain might indicate the brain metastasis of breast cancer, which is usually a late event with deleterious effects on the prognosis.^23^ In addition, previous findings^24, 25^ revealed an inverse relationship between Alzheimer’s disease and breast cancer, which might indicate a close genetic relationship between the disease and brain morphological changes mainly affecting the entorhinal cortex and hippocampus (PSC: C128_3 in **Fig. 4**).

**Figure 4:**
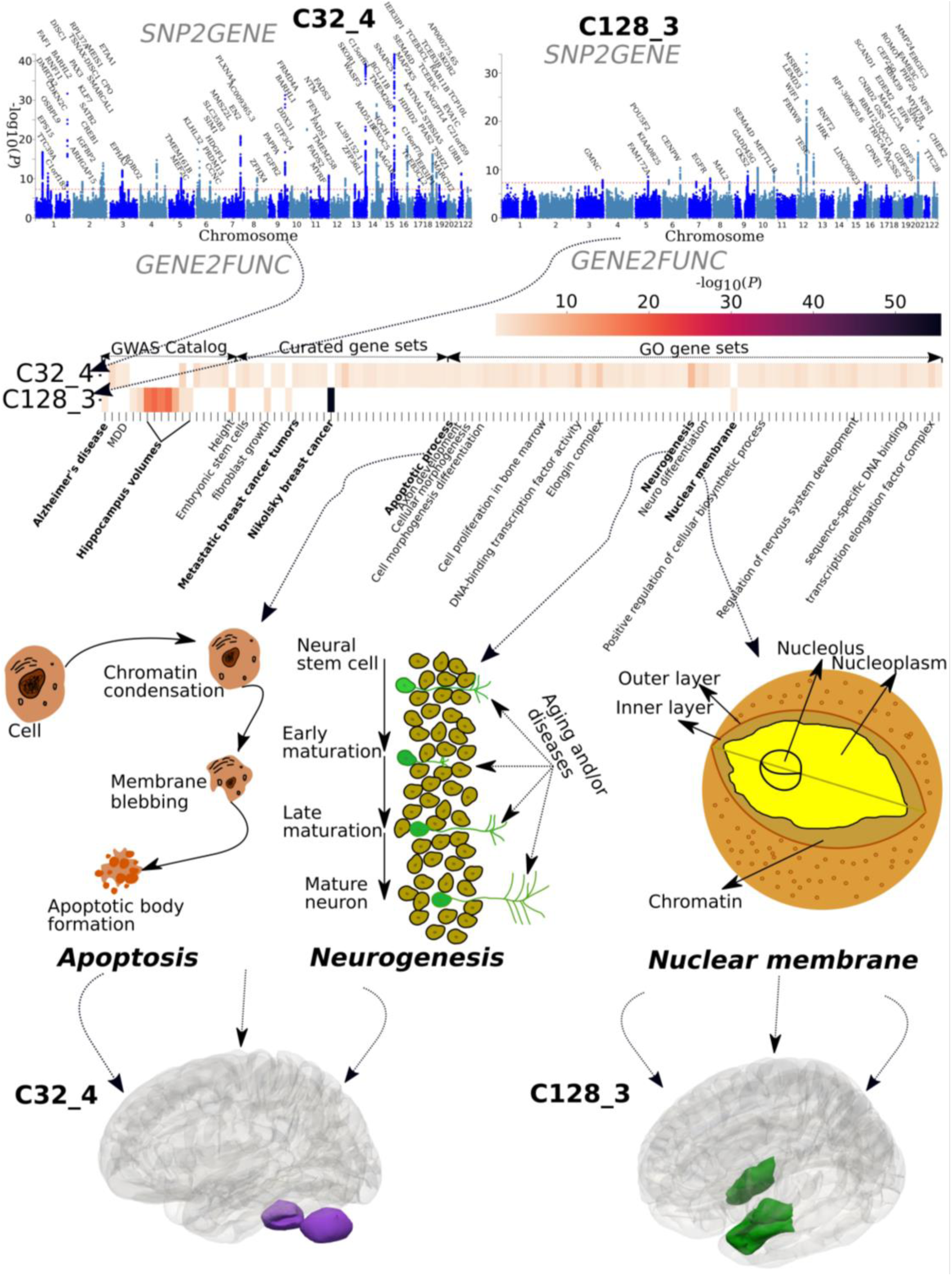
Illustrations of multiple genetic loci and pathways shaping specific patterns of structural covariance. We demonstrate how underlying genomic loci and biological pathways might influence the formation, development, and changes of two specific PSCs: the 4^th^ PSC of the C32 PSCs (C32_4) that resides in the superior part of the cerebellum and the 3^rd^ PSC of the C128 PSCs (C128_3) that includes the bilateral hippocampus and entorhinal cortex. We first performed *SNP2GENE* to annotate the mapped genes in the Manhattan plots and then ran *GENE2FUNC* for the prioritized gene set enrichment analysis (**Method 4F**). The mapped genes are input genes for prioritized gene set enrichment analyses. The heat map shows the significant gene sets from the GWAS Catalog, curated genes, and gene ontology (GO) that survived the correction for multiple comparisons. We selectively present the schematics for three pathways: apoptosis, neurogenesis, and nuclear membrane function. Several other key pathways are highlighted in bold, and the 3D maps of the two PSCs are presented.

### Illustrations of genetic loci and pathways forming two patterns of structural covariance

To illustrate how underlying genetic underpinnings might form a specific PSC, we showcased two PSCs: C32_4 for the superior cerebellum and C128_3 for the hippocampus-entorhinal cortex. The two PSCs were highly heritable and polygenic in our GWAS using the entire UKBB data (**Fig. 4**, *N*=33,541). We used the FUMA^26^ online platform to perform *SNP2GENE* for annotating the mapped genes and *GENE2FUNC* for prioritized gene set enrichment analyses (**Method 4F**). The superior cerebellum PSC was associated with genomic loci that can be mapped to 85 genes, which were enriched in many biological pathways, including psychiatric disorders, biological processes, molecular functions, and cellular components (e.g., apoptotic process, axon development, cellular morphogenesis, neurogenesis, and neuro differentiation). For example, apoptosis – the regulated cell destruction – is a complicated process that is highly involved in the development and maturation of the human brain and neurodegenerative diseases.^27^ Neurogenesis – new neuron formation – is crucial when an embryo develops and continues in specific brain regions throughout the lifespan.^28^ All significant results of this prioritized gene set enrichment analysis are presented in **Supplementary eFile 11.**

For the hippocampus-entorhinal cortex PSC, we mapped 45 genes enriched in gene sets defined from GWAS Catalog, including Alzheimer’s disease and brain volume derived from hippocampal regions. The hippocampus and medial temporal lobe have been robust hallmarks of Alzheimer’s disease.^29^ In addition, these genes were enriched in the breast cancer 20Q11 amplicon pathway^22^ and the pathway of metastatic breast cancer tumors^30^, which might indicate a specific distribution of brain metastases: the vulnerability of medial temporal lobe regions to breast cancer, ^23^ or highlight an inverse association between Alzheimer’s disease and breast cancer.^24^ Lastly, the nuclear membrane encloses the cell’s nucleus – the chromosomes reside inside – which is critical in cell formation activities related to gene expression and regulation. To further support the overlapping genetic underpinnings between this PSC and Alzheimer’s disease, we calculated the genetic correlation (*r_g_* = -0.28; p-value=0.01) using GWAS summary statistics from the hippocampus-entorhinal cortex PSC (i.e., 33,541 people of European ancestry) and a previous independent study of Alzheimer’s disease^31^ (i.e., 63,926 people of European ancestry) using LDSC.^32^ All significant results of this prioritized gene set enrichment analysis are presented in **Supplementary eFile 12.**

### Multi-scale patterns of structural covariance derive disease-related imaging signatures

We used the multi-scale PSCs from a diverse population to derive imaging signatures that reflect brain development, aging, and the effects of several brain diseases. We investigate the added value of the multi-scale PSCs as building blocks of imaging signatures for several brain diseases and risk conditions using linear support vector machines (SVM) (**Method 5**).^33^ The aim is to harness machine learning to drive a clinically interpretable metric for quantifying an individual-level risk to each disease category. To this end, we define the signatures as SPARE-X (Spatial PAtterns for REcognition) indices, where X is the disease. For instance, SPARE-AD captures the degree of expression of an imaging signature of AD-related brain atrophy, which has been shown to offer diagnostic and prognostic value in prior studies.^34^

The most discriminative indices in our samples were SPARE-AD and SPARE-MCI (**Fig. 5**, **Supplementary eTable 4a** and **eFigure 5**). C=1024 achieved the best performance for the single-scale analysis (e.g., AD vs. controls; balanced accuracy: 0.90±0.02; Cohen’s *d*: 2.50). Multi-scale representations derived imaging signatures that showed the largest effect sizes to classify the patients from the controls (**Fig. 5**) (e.g., AD vs. controls; balanced accuracy: 0.92±0.02; Cohen’s *d*: 2.61). PSCs obtained better classification performance than both AAL (e.g., AD vs. controls; balanced accuracy: 0.82±0.02; Cohen’s *d*: 1.81) and voxel-wise regional volumetric maps (RAVENS)^35^ (e.g., AD vs. controls; balanced accuracy: 0.85±0.02; Cohen’s *d*: 2.04) (**Supplementary eTable 4a** and **eFigure 5**). Our classification results were higher than previous baseline studies^36, 37^, which provided an open-source framework to objectively and reproducibly evaluate AD classification. Using the same cross-validation procedure and evaluation metric, they reported the highest balanced accuracy of 0.87±0.02 to classify AD from healthy controls. Notably, our experiments followed good practices, employed rigorous cross-validation procedures, and avoided critical methodological flaws, such as data leakage or double-dipping (refer to critical reviews on this topic elsewhere^36, 38^).

**Figure 5:**
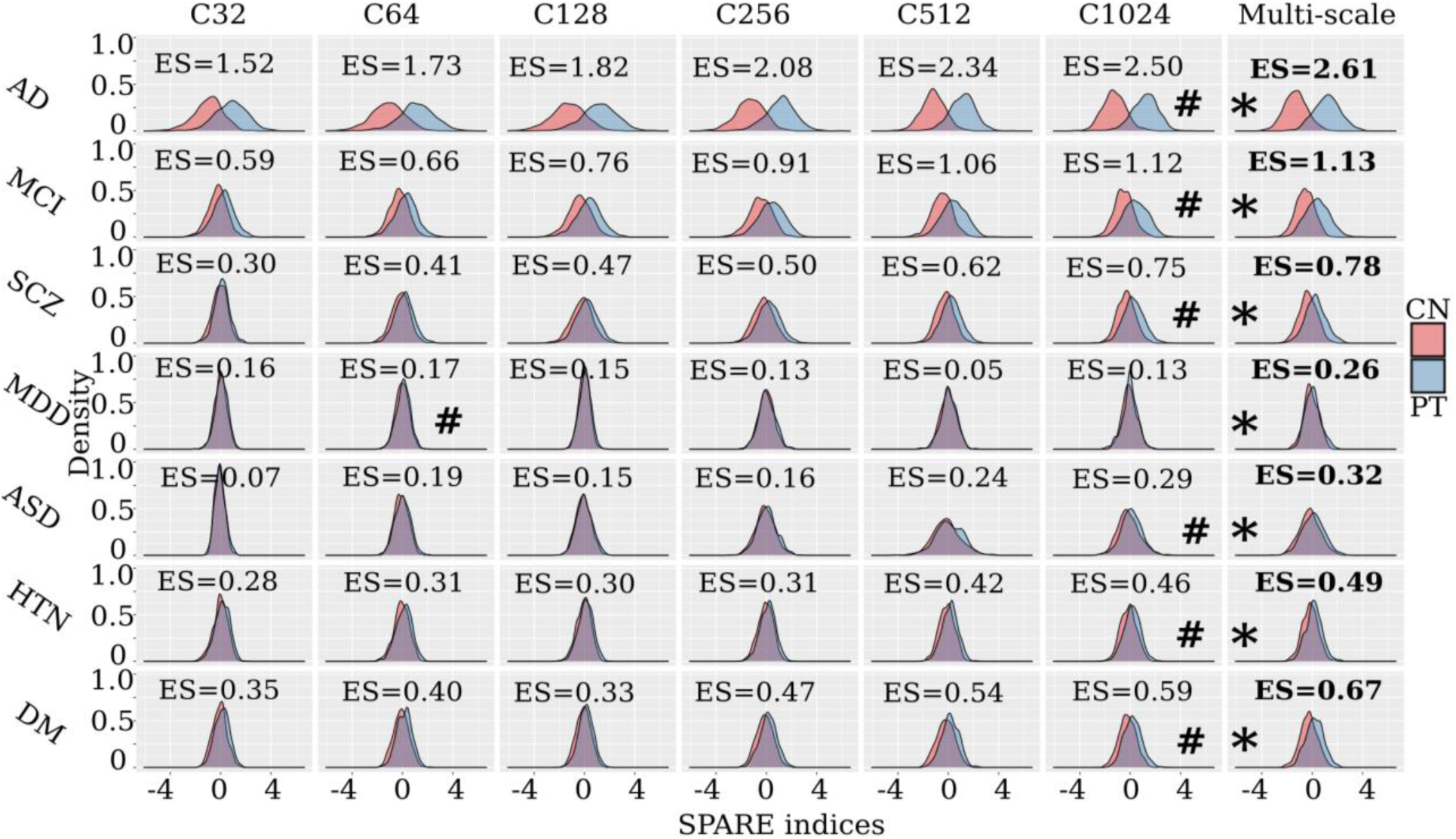
Individualized imaging signatures based on pattern analysis via machine learning. Imaging signatures (SPARE indices) of brain diseases, derived via supervised machine learning models, are more distinctive when formed from multi-scale PSCs than single-scale PSCs. The kernel density estimate plot depicts the distribution of the patient group (blue) in comparison to the healthy control group (red), reflecting the discriminative power of the diagnosis-specific SPARE (imaging signature) indices. We computed Cohen’s *d* for each SPARE index between groups to present the effect size of its discrimination power. **\*** represents the model with the largest Cohen’s *d* for each SPARE index to separate the control vs. patient groups; **#** represents the model with the best performance with single-scale PSCs. Our results demonstrate that the multi-scale PSCs generally achieve the largest discriminative effect sizes (ES) (**Supplementary eTable 4a**). As a reference, Cohen’s *d* of ≥ 0.2, ≥ 0.5, and ≥ 0.8, respectively, refer to small, moderate, and large effect sizes.

To test the robustness of these SPARE indices, we performed leave-one-site-out analyses for SPARE-AD using the combined 2003 PSCs from all scales (**Supplementary eTable 4b**). Overall, holding the ADNI data out as independent test data resulted in a lower balanced accuracy (0.88±0.02) compared to the other cases for AIBL (0.95±0.02) and PENN data (0.95±0.02). The mean balanced accuracy (0.91±0.02) aligns with the nested cross-validated results using the full sample (**Fig. 5**).

### BRIDGEPORT: bridging knowledge across patterns of structural covariance, genomics, and clinical phenotypes

We integrated our experimental results and the MuSIC atlas into the BRIDGEPORT online web portal. This online tool allows researchers to interactively browse the MuSIC atlas in 3D, query our experimental results via variants or PSCs, and download the GWAS summary statistics for further analyses. In addition, we allow users to search via conventional brain anatomical terms (e.g., the right thalamus proper) by automatically annotating traditional anatomic atlas ROIs, specifically from the MUSE atlas^39^ (**Supplementary eTable 5**), to MuSIC PSCs based on their degree of overlaps (**Supplementary eFigure 6**). Open-source software dedicated to image processing,^39^ genetic quality check protocols, MuSIC generation with sopNMF, and machine learning^36^ is also publicly available (see Code Availability for details).

## Discussion

The current study investigates patterns of structural covariance in the human brain at multiple scales from a large population of 50,699 people and, importantly, a very diverse cohort allowing us to capture patterns of structural covariance emanating from normal and abnormal brain development and aging, as well as from several brain diseases. Through extensive examination of the genetic architecture of these multi-scale PSCs, we confirmed genetic hits from previous T1-weighted MRI GWAS and, more importantly, identified 617 novel genomic loci and molecular and biological pathways that collectively influence brain morphological changes and development over the lifespan. Using a hypothesis-free, data-driven approach to first derive these PSCs using brain MRIs, we then uncovered their genetic underpinnings and further showed their potential as building blocks to predict various diseases. All experimental results and code are encapsulated and publicly available in BRIDGEPORT for dissemination: https://www.cbica.upenn.edu/bridgeport/, to enable various neuroscience studies to investigate these structural covariance patterns in diverse contexts. Together, the current study highlighted the adoption of machine learning methods in brain imaging genomics and deepened our understanding of the genetic architecture of the human brain.

Our findings reveal new insights into genetic underpinnings that influence structural covariance patterns in the human brain. Brain morphological development and changes are largely polygenic and heritable, and previous neuroimaging GWAS has not fully uncovered this genetic landscape. In contrast, genetic variants, as well as environmental, aging, and disease effects, exert pleiotropic effects in shaping morphological changes in different brain regions through specific biological pathways. The mechanisms underlying brain structural covariance are not yet fully understood. They may involve an interplay between common underlying genetic factors, shared susceptibility to aging, and various brain pathologies, which affect brain growth or degeneration in coordinated brain morphological changes.^1^ Our data-driven, multi-scale PSCs identify the hierarchical structure of the brain under the principle of structural covariance and are associated with genetic factors at different levels, including SNPs, genes, and gene set pathways. These 617 novel genomic loci, as well as those previously identified, collectively shape brain morphological changes through many key biological and molecular pathways. These pathways are widely involved in reelin signaling, apoptotic processes, axonal development, cellular morphogenesis, neurogenesis, and neuro differentiation,^27, 28^ which may collectively influence the formation of structural covariance patterns in the brain. Strikingly, pathways involved in breast cancer shared overlapping genetic underpinnings evidenced in our MAGMA-based and prioritized (*GENE2FUNC*) gene set enrichment analyses (**Fig. 3C** and **Fig. 4**), which included specific pathways involved in breast cancer and metastatic breast cancer tumors. One previous study showed that common genes might mediate breast cancer metastasis to the brain,^23^ and a later study further corroborated that the metastatic spread of breast cancer to other organs (including the brain) accelerated during sleep in both mouse and human models.^40^ We further showcased that this brain metastasis of breast cancer might be associated with specific neuropathologic processes, which were captured by PSCs data driven by Alzheimer’s disease-related neuropathology. For example, the hippocampus-entorhinal cortex PSC (C128_3, **Fig. 4**) connected the bilateral hippocampus and medial temporal lobe – the salient hallmark of Alzheimer’s disease. Our gene set enrichment analysis results further support this claim: the genes were enriched in the gene sets of Alzheimer’s disease and breast cancer (**Fig. 4**). Previous research^24, 25^ also found an inverse association between Alzheimer’s disease and breast cancer. In addition, PSCs from the cerebellum were the most genetically influenced brain regions, consistent with previous neuroimaging GWAS.^4, 5^ The cerebral cortex has been thought to largely contribute to the unique mental abilities of humans. However, the cerebellum may also be associated with a much more comprehensive range of complex cognitive functions and brain diseases than initially thought.^41^ Our results confirmed that many genetic substrates might support different molecular pathways, resulting in cerebellar functional organization, high-order functions, and dysfunctions in various brain disorders.

The current work demonstrates that appropriate machine learning analytics can be used to shed new light on brain imaging genetics. Previous neuroimaging GWAS leveraged multimodal imaging-derived phenotypes from conventional brain atlases^4, 5^ (e.g., the AAL atlas). In contrast, multi-scale PSCs are purely data-driven and likely to reflect the dynamics of underlying normal and pathological neurobiological processes giving rise to structural covariance. The diverse training sample from which the PSCs were derived, including healthy and diseased individuals of a wide age range, enriched the diversity of such neurobiological processes influencing the PSCs. In addition, modeling structural covariance at multiple scales (i.e., multi-scale PSCs) indicated that disease effects could be robustly and complementarily identified across scales (**Fig. 5**), concordant with the paradigm of multi-scale brain modeling.^13^ Imaging signatures of brain diseases, derived via supervised machine learning models, were consistently more distinctive when formed from multi-scale PSCs than single-scale PSCs. Multivariate learning techniques have gained significant prominence in neuroimaging and have recently attracted considerable attention in the domain of imaging genomics. These methods have proven valuable for analyzing complex and high-dimensional data, facilitating the exploration of relationships between imaging features and genetic factors. For instance, the MOSTest, a multivariate GWAS approach, preserves correlation structure among phenotypes via permutation on each SNP and derives a genotype vector for testing the association across all phenotypes^42^. A separate study by Soheili-Nezhad et al. demonstrated that genetic components obtained through PCA or ICA applied to neuroimaging GWAS summary statistics exhibited greater reproducibility than raw univariate GWAS effect sizes^43^. A recent study utilized a CNN-based autoencoder to discover new phenotypes and identify numerous novel genetic signals^44^. Despite the effectiveness of these multivariate approaches in GWAS, they typically conduct phenotype engineering before performing GWAS without explicitly incorporating imaging genetic associations during the modeling process. Yang et al. recently conducted a study that employed generative adversarial networks (termed GeneSGAN^45^) to integrate imaging and genetic variations within the modeling framework to address this limitation. By incorporating both modalities, their approach aimed to capture the complexity and heterogeneity of disease manifestations.

MuSIC – with the strengths of being data-driven, multi-scale, and disease-effect informative – contributes to the century-old quest for a “universal” atlas in brain cartography^46^ and is highly complementary to previously proposed brain atlases. For instance, Chen and colleagues^47^ used a semi-automated fuzzy clustering technique with MRI data from 406 twins and parcellated the cortical surface area into a genetic covariance-informative brain atlas; MuSIC was data-driven by structural covariance. Glasser and colleagues^48^ adopted a semi-automated parcellation procedure to create a multimodal cortex atlas from 210 healthy individuals. Although this method successfully integrates multimodal information from cortical folding, myelination, and functional connectivity, this semi-automatic approach requires significant resources, some with limited resolution. MuSIC allows flexible, multiple scales for delineating macroscopic brain topology; including patient samples exposes the model to sources of variability that may not be visible in healthy controls. Another pioneering endeavor is the Allen Brain Atlas project,^49^ whose overarching goals of mapping the human brain to gene expression data via existing conventional atlases, identifying local gene expression patterns across the brain in a few individuals, and deepening our understanding of the human brain’s differential genetic architecture, are complementary to ours – characterizing the global genetic architecture of the human brain, emphasizing pathogenic variability and morphological heterogeneity.

Bridging knowledge across the brain imaging, genomics, and machine learning communities is another pivotal contribution of this work. BRIDGEPORT provides a platform to lower the entry barrier for whole-brain genetic-structural analyses, foster interdisciplinary communication, and advocate for research reproducibility.^36, 50–53^ The current study demonstrates the broad applicability of this large-scale, multi-omics platform across a spectrum of neurodegenerative and neuropsychiatric diseases.

The present study has certain limitations. Firstly, the sopNMF method utilized in brain parcellation considers only imaging structural covariance and overlooks the genetic determinants contributing to forming these structural networks, as indicated by our GWAS findings. Consequently, further investigations are needed to integrate imaging and genetics into brain parcellation. Additionally, it is important to note that our GWAS analyses primarily involved participants of European ancestry. To enhance genetic findings for underrepresented ethnic groups, future studies should prioritize the inclusion of diverse ancestral backgrounds, thereby promoting a more comprehensive understanding of the genetic underpinnings across different populations.

## Methods

### Method 1: Structural covariance patterns via stochastic orthogonally projective non-negative matrix factorization

The sopNMF algorithm is a stochastic approximation built and extended based on opNMF^9, 54^. We consider a dataset of 𝑛 MR images and 𝑑 voxels per image. We represent the data as a matrix ***X*** where each column corresponds to a flattened image: 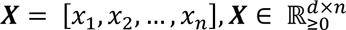. The sopNMF algorithm factorizes ***X*** into two low-rank (𝑟) matrices 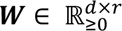 and 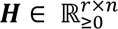 under the constraints of non-negativity and column-orthonormality. Using the Frobenius norm, the loss of this factorization problem can be formulated as

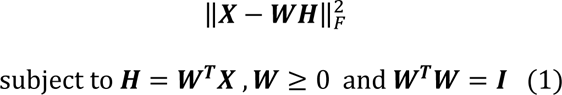

where ***I*** stands for the identity matrix. The columns 𝑤_𝑖_ ∈ ℝ^𝑑^, ‖𝑤_𝑖_‖^2^ = 1, ∀ 𝑖 ∈ {1. . 𝑟} of the so-called component matrix 𝑾 = [𝑤_1_, 𝑤_2_, …, 𝑤_𝑟_] are part-based representations promoting sparsity in data in this lower-dimensional subspace. From this perspective, the loading coefficient matrix 𝑯 represents the importance (weights) of each feature above for a given image. Instead of optimizing the non-convex problem in a batch learning paradigm (i.e., reading all images into memory) as opNMF,^9^ sopNMF subsamples the number of images at each iteration, thereby significantly reducing its memory demand by randomly drawing data batches 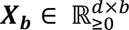 images (*b* is the batch size; *b*=32 was used in the current analyses); this is done without replacement so that all data goes through the model once (⌈𝑛/𝑏⌉). In this case, the updating rule can be rewritten as

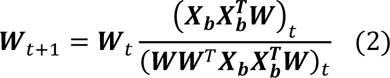

We calculate the loss on the entire dataset at the end of each epoch (i.e., the loss is incremental across all batches) with the following expression:

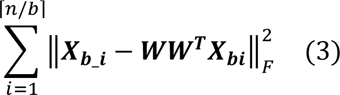

We evaluated the training loss and the sparsity of ***W*** at the end of each iteration. Moreover, early stopping was implemented to improve training efficiency and alleviate overfitting. We summarize the sopNMF algorithm in **Supplementary Algorithm 1**. An empirical comparison between sopNMF and opNMF is detailed in **Supplementary eMethod 1**.

We applied sopNMF to the training population (*N*=4000). The component matrix ***W*** was sparse after the algorithm converged with a pre-defined maximum number of epochs (100 by default) with an early stopping criterion. To build the MuSIC atlas, we clustered each voxel (row-wise) into one of the 𝑟 features/PSCs as follows:

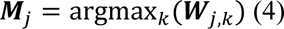

where ***M*** is a *d*-dimensional vector and 𝑗 ∈ {1. . 𝑑}. The *j*-th element of ***M*** equals *k* if 𝑾_𝑗,𝑘_is the maximum value of the *j*-th row. Intuitively, *M* indicates which of the *r* PSCs each voxel belongs to. We finally projected the vector 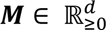 into the original image space to visualize each PSC of the MuSIC atlas (**Fig. 1**). Of note, 13 PSCs have vanished in this process for *C*=1024: all 0 for these 13 vectors.

### Method 2: Study population

We consolidated a large-scale multimodal consortium (*N*=50,699) consisting of imaging, cognition, and genetic data from 12 studies, 130 sites, and 12 countries (**Supplementary eTable 1**): the Alzheimer’s Disease Neuroimaging Initiative^55^ (ADNI) (*N*=1765); the UK Biobank^56^ (UKBB) (*N*=39,564); the Australian Imaging, Biomarker, and Lifestyle study of aging^57^ (AIBL) (*N*=830); the Biomarkers of Cognitive Decline Among Normal Individuals in the Johns Hopkins University^58^ (BIOCARD) (*N*=288); the Baltimore Longitudinal Study of Aging^59, 60^ (BLSA) (*N*=1114); the Coronary Artery Risk Development in Young Adults^61^ (CARDIA) (*N*=892); the Open Access Series of Imaging Studies^62^ (OASIS) (*N*=983), PENN (*N*=807); the Women’s Health Initiative Memory Study^63^ (WHIMS) (*N*=995), the Wisconsin Registry for Alzheimer’s Prevention^64^ (WRAP) (*N*=116); the Psychosis Heterogeneity (evaluated) via dimEnsional NeurOiMaging^65^ (PHENOM) (*N*=2125); and the Autism Brain Imaging Data Exchange^66^ (ABIDE) (*N*=1220).

We present the demographic information of the population under study in **Supplementary eTable 1**. This large-scale consortium reflects the diversity of MRI scans over different races, disease conditions, and ages over the lifespan. To be concise, we defined four populations or data sets per analysis across the paper:

- Discovery set: It consists of a multi-disease and lifespan population that includes participants from all 12 studies (*N*=32,440). Note that this population does not contain the entire UKBB population but only our first download (July 2017, *N*=21,305).
- Replication set: We held 18,259 participants from the UKBB dataset to replicate the GWAS results. We took these data from our second download of the UKBB dataset (November 2021, *N*=18,259).
- Training population: We randomly drew 250 patients (PT), including AD, MCI, SCZ, ASD, MDD, HTN (hypertension), DM (diabetes mellitus), and 250 healthy controls (CN) per decade from the discovery set, ensuring that the PT and CN groups have similar sex, study and age distributions. The resulting set of 4000 imaging data was used to generate the MuSIC atlas with the sopNMF algorithm. The rationale is to maximize variability across a balanced sample of multiple diseases or risk conditions, age, and study protocols rather than overfit the entire data by including all images in training.
- Comparison population: To validate sopNMF compared to the original opNMF algorithm, we randomly subsampled 800 participants from the training population (100 per decade for balanced CN and PT). For this scale of sample size, opNMF can load all images into memory for batch learning.^67^

All individual studies were approved by their local corresponding Institutional Review Boards (IRB). The iSTAGING and PHENOM consortia consolidated all individual imaging and clinical data; imputed genotype data were directly downloaded from the UKBB website. Data from the UKBB for this project pertains to application 35148. For iSTAGING, the IRB at the University of Pennsylvania (protocol number: 825722) reviewed the research proposal on August 31^st^, 2016, and updated it on August 31^st^, 2022. No human subjects were recruited or scanned. Existing de-identified data will be used in this mega-analysis study pooling data from 17 studies: BLSA, ADNI1, ADNI2, ADNI3, ACCORD-MIND, LookAhead, SPRINT, CARDIA, MESA, SHIP, BIOCARD, WRAP, Penn-ADC, WHIMS-MRI, AIBL, OASIS, UKBB, MESA, HANDLS. For PHENOM, the IRB at the University of Pennsylvania (protocol number: 828077) reviewed the research proposal on August 19^th^, 2017. No human subjects were recruited or scanned. Existing de-identified data will be used in this meta-analysis study pooling data from 10 studies at Penn, Ludwick-Maximmilian University of Munich, Kings College-London, University of Utrecht, University of Melbourne, University of Cantabria, University of Sao Paolo, Xijing Hospital Shaanxi, Tianjin Anning Hospital, and Institute of Mental Health Peking University.

### Method 3: Image processing and statistical harmonization

**(A) : Image processing**. Images that passed the quality check (**Supplementary eMethod 4**) were first corrected for magnetic field intensity inhomogeneity.^68^ Voxel-wise regional volumetric maps (RAVENS)^35^ for each tissue volume were then generated by using a registration method to spatially align the skull-stripped images to a template in MNI-space.^69^ We applied sopNMF to the RAVENS maps to derive MuSIC.

**(B) : Statistical harmonization of MuSIC PSCs**: We applied MuSIC to the entire population (*N*=50,699) to extract the multi-scale PSCs. Specifically, MuSIC was applied to each individual’s RAVENS gray matter map to extract the sum of brain volume in each PSC. Subsequently, the PSCs were statistically harmonized by an extensively validated approach, i.e., ComBat-GAM ^12^ (**Supplementary eMethod 3**) to account for site-related differences in the imaging data. After harmonization, the PSCs were normally distributed (skewness = 0.11±0.17, and kurtosis = 0.67±0.68) (**Supplementary eFigure 7A and B**). To alleviate the potential violation of normal distribution in downstream statistical learning, we quantile-transformed all PSCs. In agreement with the literature,^70, 71^ males were found to have larger brain volumes than females on average (**Supplementary eFigure 7C**). Overall, the Combat-GAM model slightly improved data normality across sites (**Supplementary eFigure 7E-H**). The AAL ROIs underwent the same statistical harmonization procedure.

### Method 4: Genetic analyses

Genetic analyses were restricted to the discovery and replication set from UKBB (**Method 2**). We processed the array genotyping and imputed genetic data (SNPs). The two data sets went through a “best-practice” imaging-genetics quality check (QC) protocol (**Method 4A**) and were restricted to participants of European ancestry. This resulted in 18,052 participants and 8,430,655 SNPs for the discovery set and 15,243 participants and 8,470,709 SNPs for the replication set.

We reperformed the genetic QC and genetic analyses for the combined populations for BRIDGEPORT, resulting in 33,541 participants and 8,469,833 SNPs. **Method 4G** details the correction for multiple comparisons throughout our analyses.

**(A) : Genetic data quality check protocol**. First, we excluded related individuals (up to 2^nd^- degree) from the complete UKBB sample (*N*=488,377) using the KING software for family relationship inference.^72^ We then removed duplicated variants from all 22 autosomal chromosomes. We also excluded individuals for whom either imaging or genetic data were not available. Individuals whose genetically identified sex did not match their self-acknowledged sex were removed. Other excluding criteria were: i) individuals with more than 3% of missing genotypes; ii) variants with minor allele frequency (MAF) of less than 1%; iii) variants with larger than 3% missing genotyping rate; iv) variants that failed the Hardy-Weinberg test at 1×10^-10^. To adjust for population stratification,^73^ we derived the first 40 genetic principle components (PC) using the FlashPCA software^74^. The genetic pipeline was also described elsewhere^75^.

**(B) : Heritability estimates and genome-wide association analysis**. We estimated the SNP-based heritability explained by all autosomal genetic variants using GCTA-GREML.^76^ We adjusted for confounders of age (at imaging), age-squared, sex, age-sex interaction, age-squared- sex interaction, ICV, and the first 40 genetic principal components (PC), guided by a previous neuroimaging GWAS^4^. In addition, Elliot et al.^5^ investigated more than 200 confounders in another study. Therefore, our sensitivity analyses included four additional imaging-related covariates (i.e., brain positions and head motion). One-side likelihood ratio tests were performed to derive the heritability estimates. In GWAS, we performed a linear regression for each PSC and included the same covariates as in the heritability estimates using PLINK.^77^

**(C) : Identification of novel genomic loci**. Using PLINK, we clumped the GWAS summary statistics based on their linkage disequilibrium to identify the genomic loci (see **Supplementary eMethod 5** for the definition of the index, candidate, independent significant, lead SNP, and genomic locus). In particular, the threshold for significance was set to 5×10^-8^ (*clump-p1*) for the index SNPs and 0.05 (*clump-p2*) for the **candidate SNPs**. The threshold for linkage disequilibrium-based clumping was set to 0.60 (*clump-r2*) for **independent significant SNPs** and 0.10 for lead SNPs. The linkage disequilibrium physical-distance threshold was 250 kilobases (*clump-kb*). **Genomic loci** consider linkage disequilibrium (within 250 kilobases) when interpreting the association results. The GWASRAPIDD^78^ package (version: 0.99.14) was then used to query the genomic loci for any previously-reported associations with clinical phenotypes documented in the NHGRI-EBI GWAS Catalog^15^ (p-value < 1.0×10^-5^, default inclusion value of GWAS Catalog). We defined a genomic locus as **novel** when it was not present in GWAS Catalog (query date: April 5^th^, 2023).

**(D) : Gene-level associations with MAGMA**. We performed gene-level association analysis using MAGMA.^17^ First, gene annotation was performed to map the SNPs (reference variant location from Phase 3 of 1,000 Genomes for European ancestry) to genes (human genome Build 37) according to their physical positions. The second step was to perform the gene analysis based on the GWAS summary statistics to obtain gene-level p-values between the pairwise 2003 PSCs and the 18,097 protein-encoding genes containing valid SNPs.

**(E) : Hypothesis-free gene set enrichment analysis with MAGMA**. Using the gene-level association p-values, we performed gene set enrichment analysis using MAGMA. Gene sets were obtained from Molecular Signatures Database (MsigDB, v7.5.1),^79^ including 6366 curated gene sets and 10,402 Gene Ontology (GO) terms. All other parameters were set by default for MAGMA. This hypothesis-free analysis resulted in a more stringent correction for multiple comparisons (i.e., by the total number of tested genes and PSCs) than the FUMA-prioritized gene set enrichment analysis (see below F).

**(F) : FUMA analyses for the illustrations of specific PSCs**. In *SNP2GENE*, three different methods were used to map the SNPs to genes. First, positional mapping maps SNPs to genes if the SNPs are physically located inside a gene (a 10 kb window by default). Second, expression quantitative trait loci (eQTL) mapping maps SNPs to genes showing a significant eQTL association. Lastly, chromatin interaction mapping maps SNPs to genes when there is a significant chromatin interaction between the disease-associated regions and nearby or distant genes.^26^ In addition, *GENE2FUNC* studies the expression of prioritized genes and tests for the enrichment of the set of genes in pre-defined pathways. We used the mapped genes as prioritized genes. The background genes were specified as all genes in FUMA, and all other parameters were set by default. We only reported gene sets with adjusted p-value < 0.05.

**(G) : Correction for multiple comparisons**. We practiced a conservative procedure to control for the multiple comparisons. In the case of GWAS, we chose the default genome-wide significant threshold (5.0×10^-8^, and 0.05 for all other analyses) and independently adjusted for multiple comparisons (Bonferroni methods) at each scale by the number of PSCs. We corrected the p-values for the number of phenotypes (*N*=6) for genetic correlation analyses. We adjusted the p-values for the number of PSCs at each scale for heritability estimates. For gene analyses, we controlled for both the number of PSCs at each scale and the number of genes. We adopted these strategies per analysis to correct the multiple comparisons because PSCs of different scales are likely hierarchical and correlated – avoiding the potential of “overcorrection”.

**(H) : Replication analysis for genome-wide association studies**. We performed GWAS by fitting the same linear regressing models as the discovery set. Also, following the same procedure for consistency, we corrected the multiple comparisons using the Bonferroni method. We corrected it for the number of genomic loci (*N*=915) found in the discovery set with a nominal p-value of 0.05, which thereby resulted in a stringent test with an equivalent p-value threshold of 3.1×10^-5^ (i.e., (-log_10_[p-value] = 4.27). We performed a replication for the 915 genomic loci, but, in reality, SNPs in linkage disequilibrium with the genomic loci are likely highly significant.

### Method 5: Pattern analysis via machine learning for individualized imaging signatures

SPARE-AD captures the degree of expression of an imaging signature of AD, and prior studies have shown its diagnostic and prognostic values.^34^ Here, we extended the concept of the SPARE imaging signature to multiple diseases (SPARE-X, X represents disease diagnoses). Following our reproducible open-source framework^37^, we performed nested cross-validation (**Supplementary eMethod 6**) for the machine learning models and derived imaging signatures to quantify individualized disease vulnerability.

**SPARE indices**. MuSIC PSCs were fit into a linear support vector machine (SVM) to derive SPARE-AD, MCI, SCZ, DM, HTN, MDD, and ASD. Specifically, the SVM aims to classify the patient group (e.g., AD) from the control group and outputs a continuous variable (i.e., the SPARE indices), which indicates the proximity of each participant to the hyperplane in either the patient or control space. We compared the classification performance using different sets of features: i) the single-scale PSC from 32 to 1024, ii) the multi-scale PSCs by combining all features (with and without feature selections embedded in the CV); iii) the ROIs from the AAL atlas; and iv) voxel-wise RAVENS maps. The samples selected for each task are presented in **Supplementary eTable 2**.

No statistical methods were used to predetermine the sample size. The experiments were not randomized, and the investigators were not blinded to allocation during experiments and outcome assessment.

## Data Availability

The GWAS summary statistics corresponding to this study are publicly available on the BRIDGEPORT web portal (https://www.cbica.upenn.edu/bridgeport/) and the MEDICINE web portal (http://labs.loni.usc.edu/medicine/). The GWAS summary statistics used in the genetic correlation analyses were fetched from the GWAS Catalog platform (https://www.ebi.ac.uk/gwas), although each study provided the original links; The GWAS Catalog platform was used to query if the SNPs identified by MuSIC were previously reported.

## Code Availability

The software and resources used in this study are all publicly available:

- sopNMF: https://pypi.org/project/sopnmf/, MuSIC, and sopNMF (developed for this study)
- BRIDGEPORT: https://www.cbica.upenn.edu/bridgeport/, (developed for this study)
- MLNI: https://pypi.org/project/mlni/, machine learning (developed for this study)
- MUSE: https://www.med.upenn.edu/sbia/muse.html, image preprocessing
- PLINK: https://www.cog-genomics.org/plink/, GWAS
- GCTA: https://yanglab.westlake.edu.cn/software/gcta/#Overview, heritability estimates
- LDSC: https://github.com/bulik/ldsc, genetic correlation estimates
- MAGMA: https://ctg.cncr.nl/software/magma, gene analysis
- GWASRAPIDD: https://rmagno.eu/gwasrapidd/articles/gwasrapidd.html, GWAS Catalog query
- MsigDB: https://www.gsea-msigdb.org/gsea/msigdb/, gene sets database

## Authors’ contributions

Dr. Wen takes full responsibility for the integrity of the data and the accuracy of the data analysis.

*Study concept and design*: Wen, Davatzikos

*Acquisition, analysis, or interpretation of data*: Wen, Nasrallah, Davatzikos, Abdulkadi, Satterthwaite, Dazzan, Kahn, Schnack, Zanetti, Meisenzahl, Busatto, Crespo-Facorro, Pantelis, Wood, Zhuo, Koutsouleris, Wittfeld, Grabe, Marcus, LaMontagne, Heckbert, Austin, Launer, Espeland, Masters, Maruff, Fripp, Johnson, Morris, Albert, Resnick, Saykin, Thompson, Li, Wolf, Raquel Gur, Ruben Gur, Shinohara, Tosun-Turgut, Fan, Shou, Erus, Wolk

*Drafting of the manuscript*: Wen, Nasrallah, Davatzikos

*Critical revision of the manuscript for important intellectual content*: all authors

*Statistical and genetic analysis*: Wen

## Acknowledgments

The iSTAGING consortium is a multi-institutional effort funded by NIA by RF1 AG054409. The Baltimore Longitudinal Study of Aging neuroimaging study is funded by the Intramural Research Program, National Institute on Aging, National Institutes of Health and by HHSN271201600059C. The BIOCARD study is in part supported by NIH grant U19-AG033655. The PHENOM study is funded by NIA grant R01MH112070 and by the PRONIA project as funded by the European Union 7th Framework Program grant 602152. Other supporting funds are 5U01AG068057, 1U24AG074855, and S10OD023495. Data were provided [in part] by OASIS OASIS-3: Principal Investigators: T. Benzinger, D. Marcus, J. Morris; NIH P50 AG00561, P30 NS09857781, P01 AG026276, P01 AG003991, R01 AG043434, UL1 TR000448, R01 EB009352. AV-45 doses were provided by Avid Radiopharmaceuticals, a wholly owned subsidiary of Eli Lilly. OC has received funding from the French government under management of Agence Nationale de la Recherche as part of the “Investissements d’avenir” program, reference ANR-19-P3IA-0001 (PRAIRIE 3IA Institute) and reference ANR-10-IAIHU-06 (Agence Nationale de la Recherche-10-IA Institut Hospitalo-Universitaire-6). This research has been conducted using the UK Biobank Resource under Application Number 35148. Data used in preparation of this article were in part obtained from the Alzheimer’s Disease Neuroimaging Initiative (ADNI) database (adni.loni.usc.edu). As such, the investigators within the ADNI contributed to the design and implementation of ADNI and/or provided data but did not participate in the analysis or writing of this report. A complete listing of ADNI investigators can be found at: http://adni.loni.usc.edu/wpcontent/uploads/how_to_apply/ADNI_Acknowledgement_List.pdf. ADNI is funded by the National Institute on Aging, the National Institute of Biomedical Imaging and Bioengineering, and through generous contributions from the following: AbbVie, Alzheimer’s Association; Alzheimer’s Drug Discovery Foundation; Araclon Biotech; BioClinica, Inc.; Biogen; Bristol-Myers Squibb Company; CereSpir, Inc.; Cogstate; Eisai Inc.; Elan Pharmaceuticals, Inc.; Eli Lilly and Company; EuroImmun; F. Hoffmann-La Roche Ltd and its affiliated company Genentech, Inc.; Fujirebio; GE Healthcare; IXICO Ltd.; Janssen Alzheimer Immunotherapy Research & Development, LLC.; Johnson & Johnson Pharmaceutical Research & Development LLC.; Lumosity; Lundbeck; Merck & Co., Inc.; Meso Scale Diagnostics, LLC.; NeuroRx Research; Neurotrack Technologies; Novartis Pharmaceuticals Corporation; Pfizer Inc.; Piramal Imaging; Servier; Takeda Pharmaceutical Company; and Transition Therapeutics. The Canadian Institutes of Health Research is providing funds to support ADNI clinical sites in Canada. Private sector contributions are facilitated by the Foundation for the National Institutes of Health (www.fnih.org). The grantee organization is the Northern California Institute for Research and Education, and the study is coordinated by the Alzheimer’s Therapeutic Research Institute at the University of Southern California. ADNI data are disseminated by the Laboratory for Neuro Imaging at the University of Southern California. Dr. Wen and Dr. Davatzikos had full access to all the data in the study. They took responsibility for the integrity of the data and the accuracy of the data analysis.

## eMethod 1: Empirical validation of sopNMF

For the empirical validation of sopNMF, the comparison population (**Method 1** in the main manuscript) was used so that the machine’s memory could be sufficient to read the entire data for opNMF. For sopNMF, different choices of batch size (i.e., BS=32, 64, 128, and 256) were tested. We hypothesized that sopNMF could approximate the optima of opNMF during optimization, i.e., resulting in similar parts-based representation, training loss, and sparsity. TensorboardX was embedded into the sopNMF framework to monitor the training process dynamically. All experiments were performed on an Ubuntu machine with a maximum RAM of 32 GB and 8 CPUs. The predefined maximum number of epochs for all experiments is 50,000, and the tolerance of early stopping criteria is 100 epochs based on the training loss.

We qualitatively compared the extracted PSCs and quantitatively for the training loss, the sparsity of the component matrix ***W***, and the memory consumption for *C*=20 (number of PSCs). The 20 PSCs were spatially consistent between opNMF and sopNMF, despite that some regions were decomposed into different PSCs (i.e., the white ellipse in **eFig. 1A**). For the training loss, opNMF obtained the lowest loss (1.103 x 10^6^), and the loss of sopNMF were 1.107 x10^6^, 1.108 x10^6^, 1.111 x10^6^ and 1.210 x10^6^ for BS =256, 128, 64, and 32, respectively (**eFig. 1D**). For the sparsity of the component matrix, all models obtained comparable results (sparsity ≈ 0.83, **eFig. 1E**). The estimated memory consumptions during the training process were 28.65, 4.02, 3.81, 2.60, 1.47 GB for opNMF and sopNMF (BS =256, 128, 64, and 32), respectively (**Fig. e1F**).

## eMethod 2: Reproducibility index

We proposed a reproducibility index (RI) to test the reproducibility of sopNMF for brain parcellation:

- We used the Hungarian match algorithm^80^ to match the pairs of PSCs between two splits under the specific condition that maximizes the similarity (i.e., minimizes the cost of workers/jobs in its original formulation).
- For each pair of PSCs, we calculated the inner product of the vectors (𝑅^𝑑^), referred to as RI. This index takes values between [0, 1], with higher values indicating higher reproducibility.
- For each scale *C*, we presented the mean/standard deviation of the RIs for all PSCs.

## eMethod 3: Inter-site image harmonization

We used an extensively validated statistical harmonization approach, i.e., ComBat-GAM,^12^ to harmonize the extracted multi-scale PSCs. This method estimates the variability in volumetric measures due to differences in site/cohort-specific imaging protocols based on variances observed within and across control groups while preserving normal variances due to age, sex, and intracranial volume (ICV) differences. The model was initially trained on the discovery set and then applied to the replication set.

## eMethod 4: Quality check of the image processing pipeline

Raw T1-weighted MRIs were first quality checked (QC) for motion, image artifacts, or restricted field-of-view. Another QC was performed: First, the images were examined by manually evaluating for pipeline failures (e.g., poor brain extraction, tissue segmentation, and registration errors). Furthermore, a second step automatically flagged images based on outlying values of quantified metrics (i.e., PSC values); those flagged images were re-evaluated.

## eMethod 5: Definition of the index, candidate, independent significant, and lead SNP and genomic locus

### Index SNP

They are defined as SNPs with a p-value threshold ≤ 5e-8 (*clump-p1*) from GWAS summary statistics.

### Independent significant SNP

They are defined as the index SNPs, which are independent of each other (not in linkage disequilibrium) with *r^2^* ≤ 0.6 (*clump-r2*) within 250 kilobases (non-overlapping, *clump-kb*) away from each other.

### Lead SNP and genomic loci

They are defined as the independent significant SNPs, which are independent of each other with a more stringent *r^2^* ≤ 0.1 (*clump-r2*) within 250 kilobases (non-overlapping, *clump-kb*) away from each other. Each of these clumps is defined as a *genomic locus*.

### Candidate SNP

With each genomic locus, candidate SNPs are defined as the SNPs whose association p-values are smaller than 0.05 (*clump-p2*). The definitions followed instructions from FUMA^26^ and Plink^77^ software.

## eMethod 6: Cross-validation procedure for PAML

Nested cross-validation was adopted for all tasks following the good-practice guidelines proposed in our previous works^36, 37, 53^. In particular, an outer loop was used to evaluate the task performance (250 repetitions of random hold-out splits with 80% of data for training). In contrast, an inner loop focused on tuning the hyperparameters (10-fold splits). We computed the balanced accuracy (BA) to evaluate the classification tasks. We calculated the effect size (Cohen’s *d*) and p-value for each SPARE index to quantify its discriminative power.

---

**eFigure 1:**
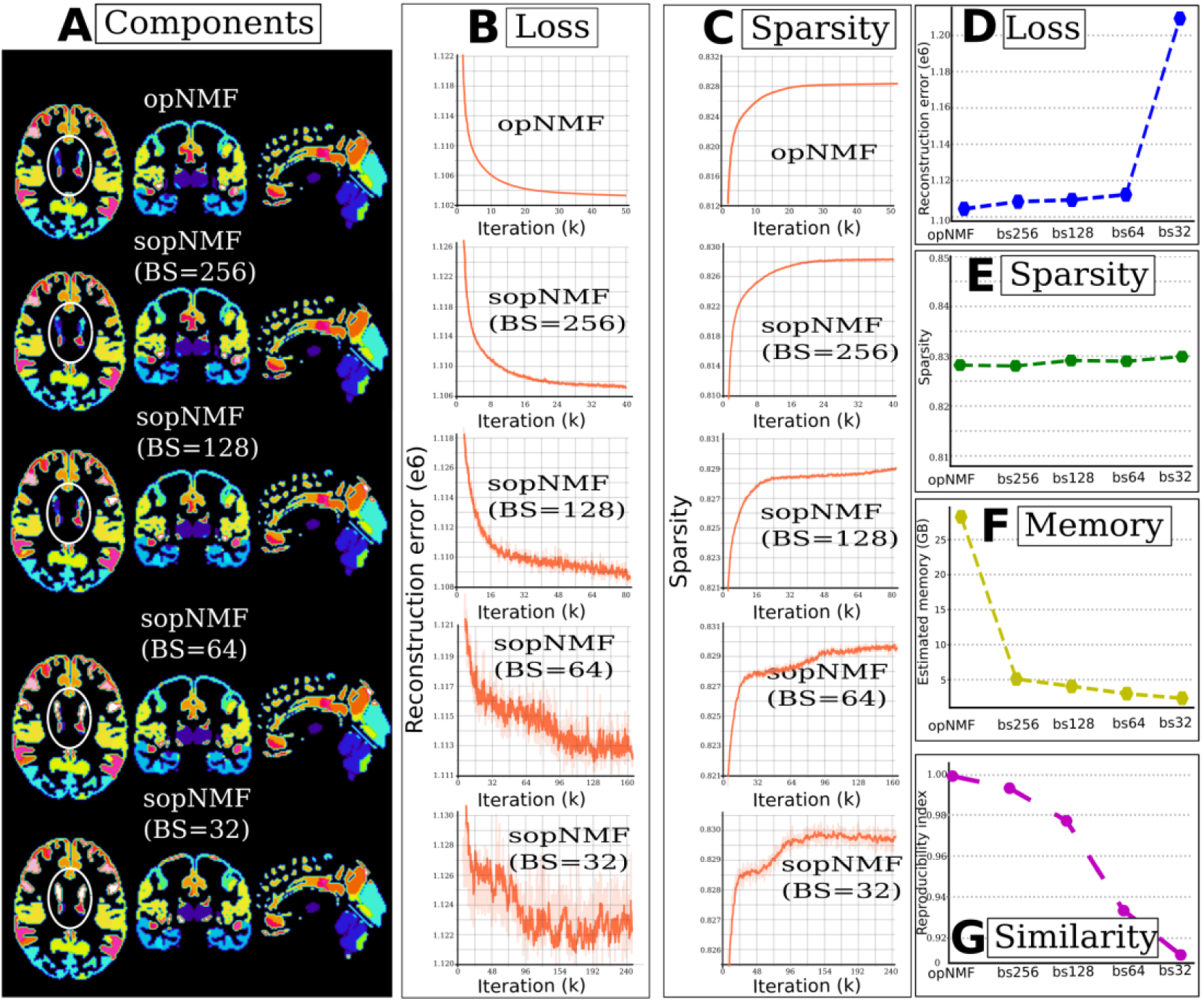
Comparison between opNMF and sopNMF. (**A**) Qualitative evaluation: The extracted components are shown in the original image space, with each PSC displayed in a distinct color. The white ellipse indicates the region where the models diverge. Quantitative evaluation: training loss (**B**, **D**) and sparsity (**C**, **E**) demonstrated similar patterns between models, except that batch size (BS) = 32 had a larger loss than the other models. Comparing the estimated memory consumption during training across models shows significant advantages for all sopNMF models compared to opNMF.

**eFigure 2:**
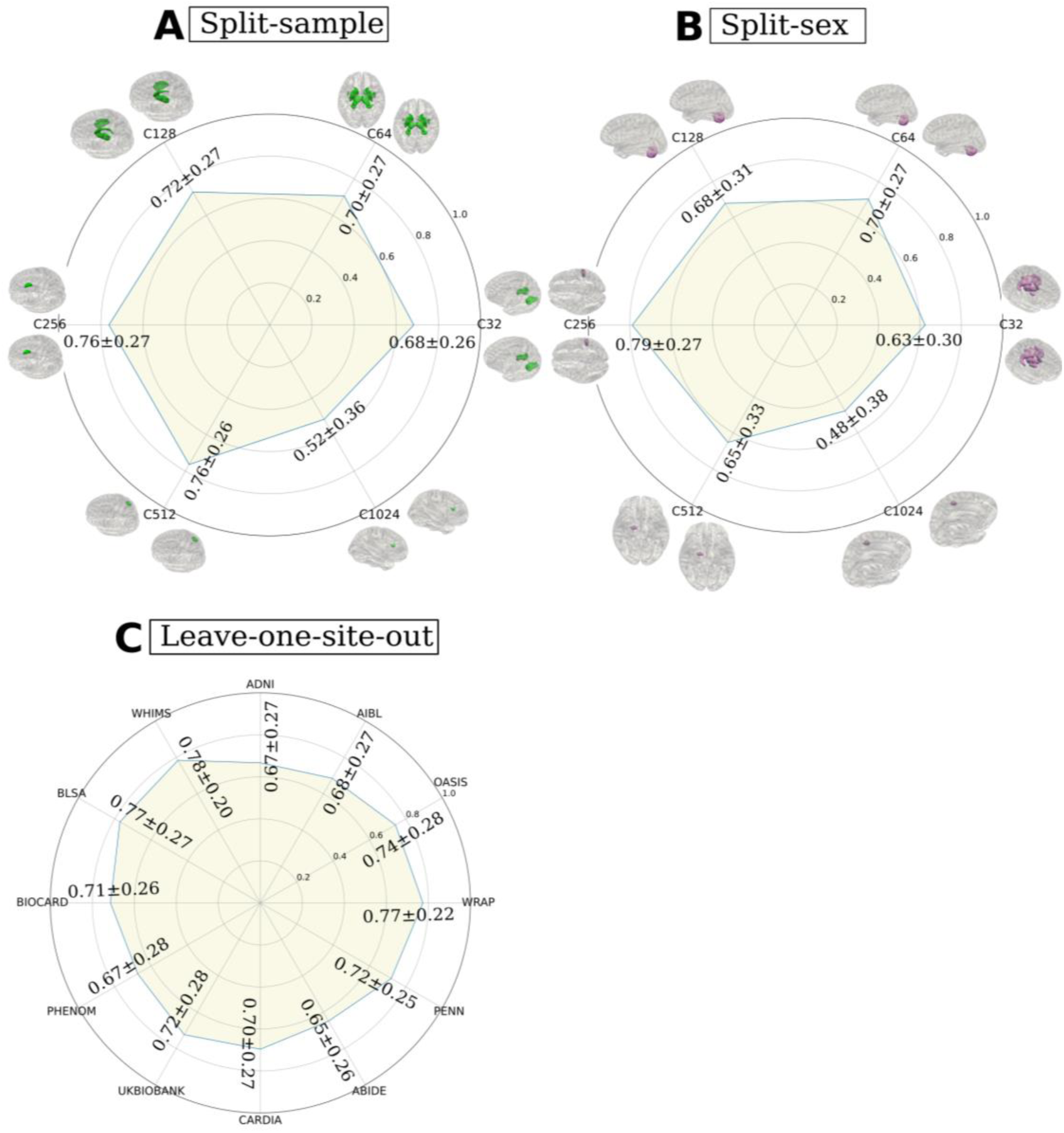
Reproducibility of the sopNMF brain parcellation. In general, sopNMF demonstrated high reproducibility under various conditions. For each brain PSC, the reproducibility index (RI) was calculated (**Supplementary eMethod 2**). (**A**) Split-sample analyses, where the training population (*N*=4000) was randomly split into two halves while maintaining similar age, sex, and site distribution between groups. (**B**) Split-sex analyses, where the training population was divided into males and females. Colored PSCs on the brain template illustrate the same PSC independently derived from the two splits. (**C**) Leave-one-site-out analyses for C32 PSCs., where the training populations excluding participants from each site (BIOCARD, ADNI, WARP, AIBL, ABIDE, BLSA, OASIS, CARDIA, PHENOM, PENN, UKBB, and WHIMS) were independently trained with sopNMF. The RI indices were compared to the sopNMF results using the full training sample (N=4000).

**eFigure 3:**
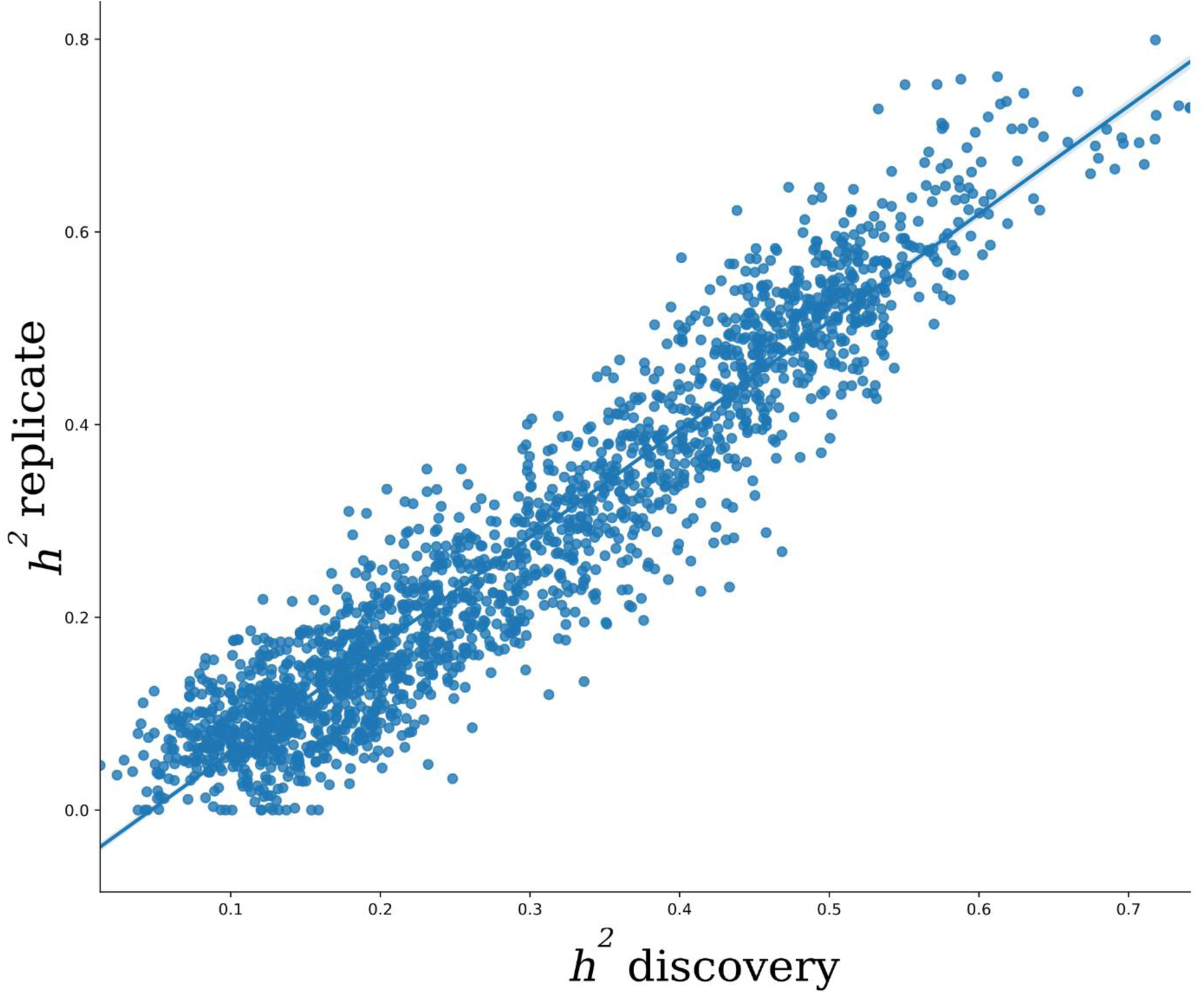
Scatter plot for the *h*^2^ estimates from the discovery and replication sets. The SNP-based heritability was estimated independently for the discovery set (*N*=18,052) and replication set (*N*=15,243). In particular, the two estimates were highly correlated (*r* = 0.94, p-value < 10^-6^), demonstrating a highly similar genetic architecture across different sets of UKBB data.

**eFigure 4:**
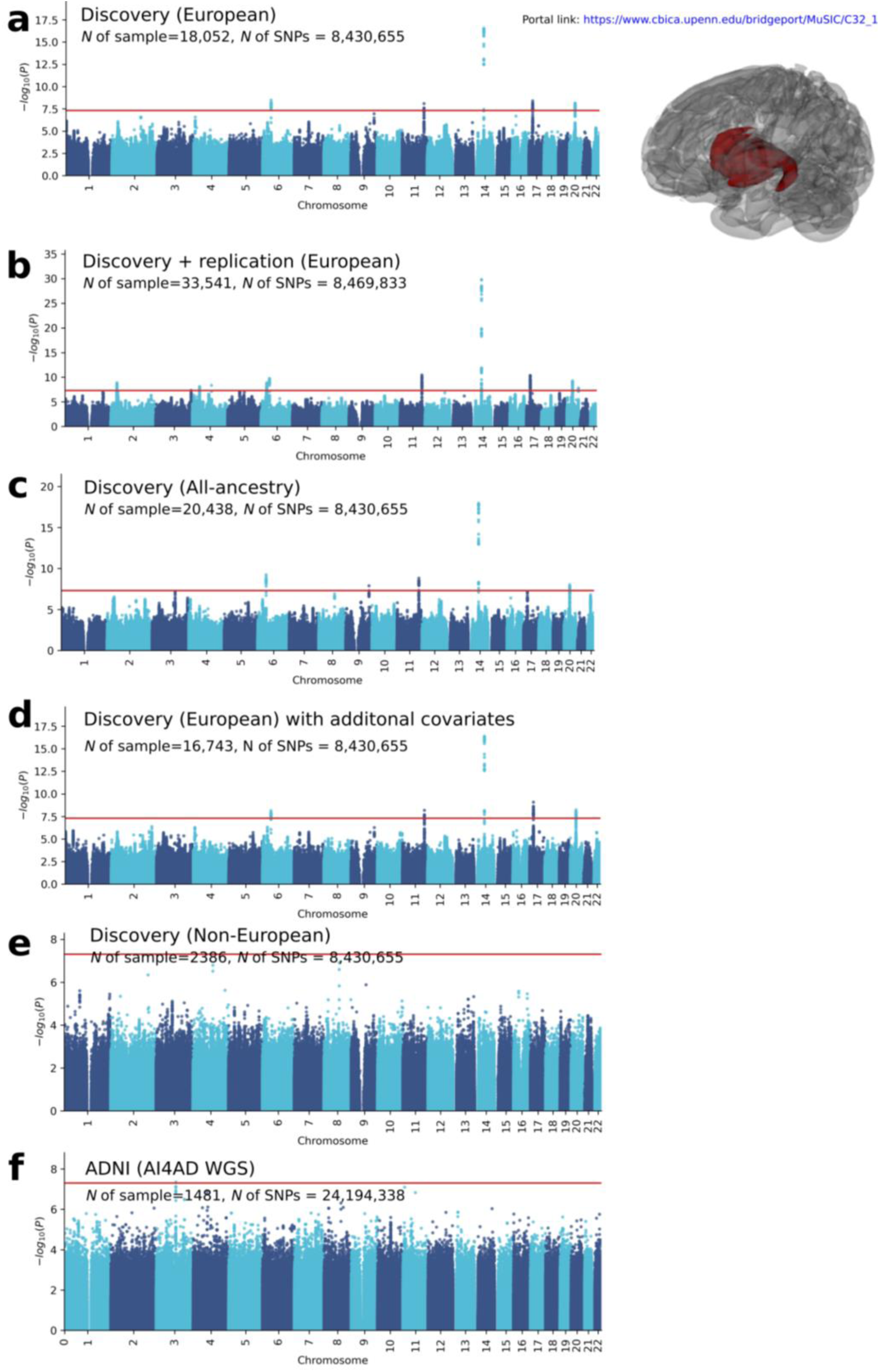
Sensitivity check for the GWAS results using the discovery set in UKBB. **A**) The GWAS results for participants with European ancestry in the discovery set. **B**) The GWAS results for participants with European ancestry in the discovery and replication sets. **C**) The GWAS results for participants with all different ancestries in the discovery set. **D**) The GWAS results for participants with European ancestry in the discovery set by adding four additional imaging-related covariates. **E**) The GWAS results for participants with non-European ancestry in the discovery set. **F**) The GWAS results for participants with the independent ADNI WGS data.

**eFigure 5:**
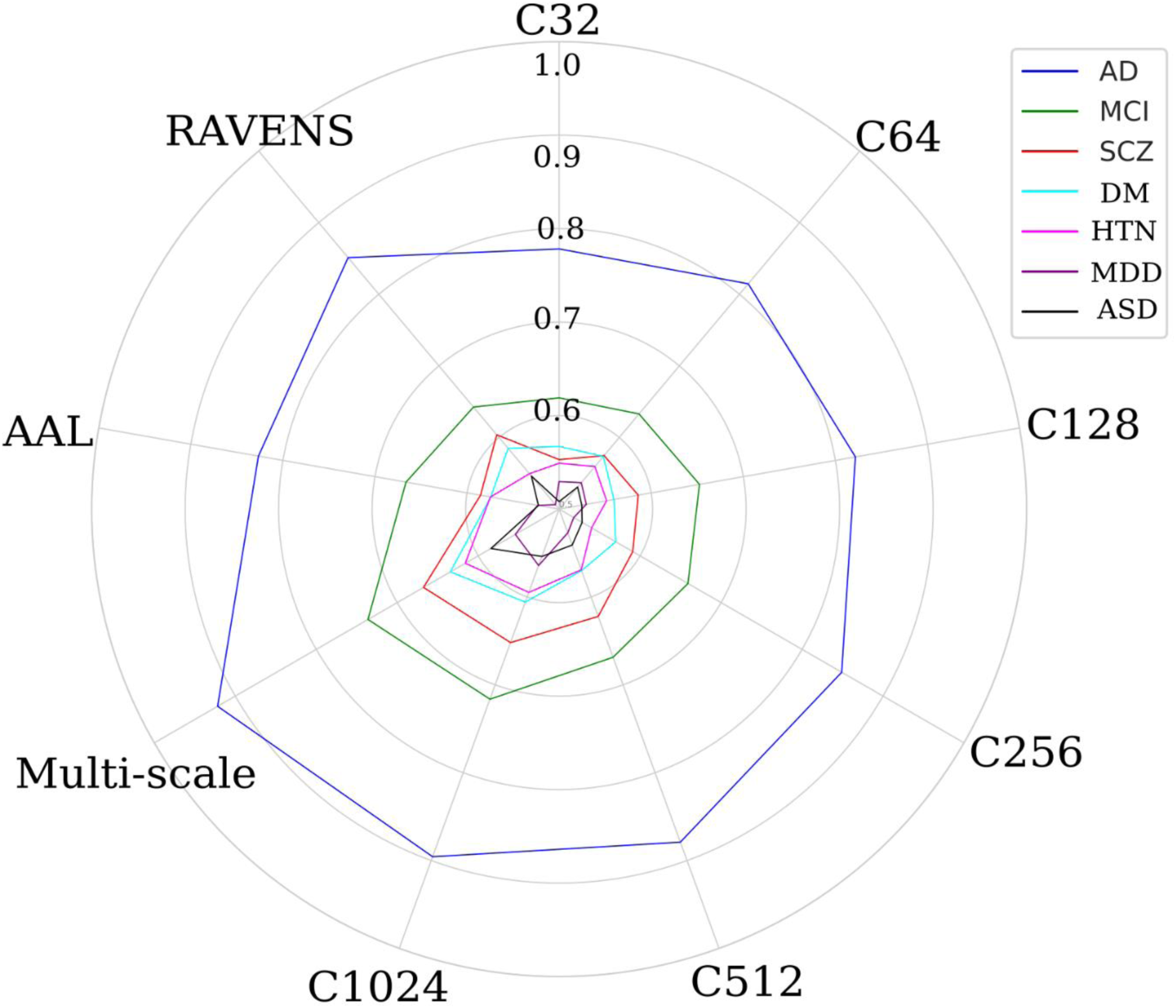
Machine learning performance for disease classification. Balanced accuracy (BA) for each classification task using different features from multi-scale MuSIC, AAL, and RAVENS (higher score better). Details are presented in **eTable 4**.

**eFigure 6:**
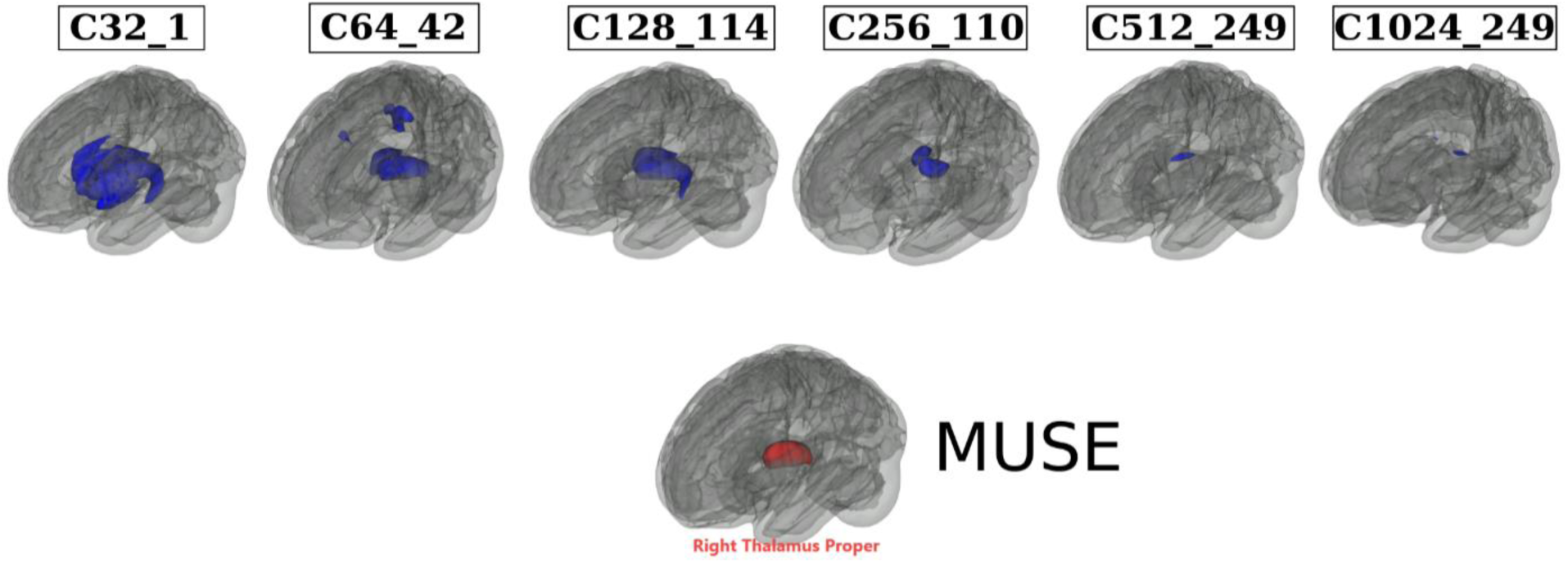
Annotation of MUSE ROIs to MuSIC PSCs based on the overlap index. We automatically annotated the 119 MUSE GM PSCs to the MuSIC atlases at all six scales (*C*=32, 64, 128, 256, 512, and 1024). To this end, we calculated an overlap index (OI) to quantify the spatial overlaps between MUSE and MuSIC. For instance, for each MUSE PSC (**eTable 5**) vs. each of the 32 PSCs of MuSIC at *C*=32 scale, the OI equals the proportion of the number of overlap voxels and the total number of voxels in the MUSE PSC. Here we illustrate by mapping the right thalamus of MUSE to all 6 MuSIC atlases. The highest OIs are 0.82, 0.70, 0.86, 0.30, 0.09, 0.05 for C32_1, C64_42, C128_114, C256_110, C512_249, and C1024_249 PSCs. This functionality is available in BRIDGEPORT: https://www.cbica.upenn.edu/bridgeport/MUSE/Right%20Thalamus%20Proper

**eFigure 7:**
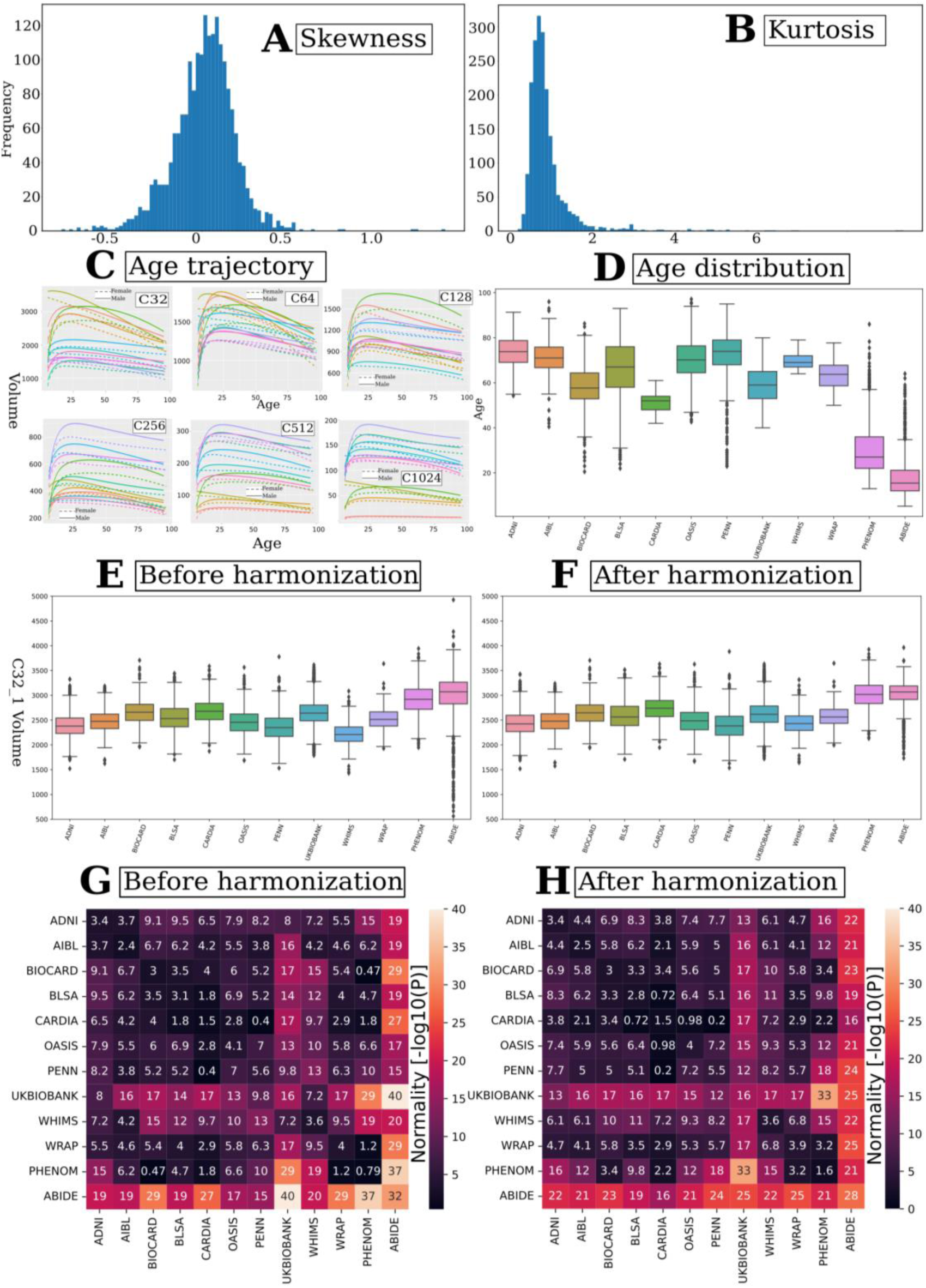
Summary statistics of the multi-scale PSCs of MuSIC. Multi-scale PSCs show considerable normal distributions, i.e., symmetrical distribution (**A**) with a low kurtosis (**B**). Moreover, we fit the Generalized Additive Model for Location, Scale, and Shape (GAMLSS)^81^ model (fractional polynomials with 2 degrees) to each PSC to delineate the age trajectory over the lifespan in males (solid lines) and females (dotted lines), respectively (**C**). For visualization purposes, we selectively display the first 10 PSCs from each scale of the MuSIC atlases. In general, males have larger brain volumes than females. For **D-F**, we selectively showed the distribution of age (**D**) and the distribution of PSC volume before harmonization (**E**) and after harmonization (**F**) for C32_1 within each site in the discovery set. For **G** and **H**, we tested the normality of the PSC volume (C32_1) from each pair of sites using the Shapiro-Wilk test (*scipy.stats.shapiro* function) in the discovery set before (**G**) harmonization and after harmonization (**H**). A higher -log_10_(P) indicates the data are less likely to be normally distributed. As a general trend, our statistical harmonization techniques demonstrated a slight improvement in the normality of the data. Additionally, we consistently applied normality transformations to all statistical analyses, including GWAS, to mitigate any non-normality.

**eTable 1.** Study cohort characteristics. The current study consists of two main populations/sets: the discovery set (*N*=32,440, including participants from the first download of the UKBB data) and the replication set (*N*=18,259, the second download of the UKBB data). To train the sopNMF model for MuSIC, we selected 250 patients (PT) and 250 healthy controls (CN) for each decade of the discovery set, resulting in 4000 participants in total, referred to as the training population. Age ranges from 5 to 97 years and is shown with mean and standard deviation. Sex is displayed with the number and percentage of female participants. Data was collected from 12 studies, 130 sites, and 12 countries. The number of sites (country) per study is detailed as follows:

| Study | *N*<br>(50,699) | Age<br>(5-97<br>year) | Sex<br>(female/%<br>) | CN<sup>b</sup> | AD | MCI | SCZ | ASD | MDD | DM | HTN |
| --- | --- | --- | --- | --- | --- | --- | --- | --- | --- | --- | --- |
| **Discovery set** | 32,440 | 60.04±14.87 | 16,868/52 | 24,980 | 954 | 1288 | 1094 | 597 | 1476 | 1093 | 958 |
| ADNI | 1765 | 73.66±7.19 | 798/45 | 297 | 343 | 875 | NA | NA | NA | NA | 250 |
| UKBB<sup>a</sup> | 21,305 | 62.58±7.48 | 10,101/53 | 18,735 | 1 | NA | NA | NA | 1476 | 1093 | NA |
| AIBL | 830 | 71.36±6.78 | 471/57 | 625 | 86 | 115 | NA | NA | NA | NA | 4 |
| BIOCARD | 288 | 58.15±10.54 | 115/60 | 283 | 1 | 4 | NA | NA | NA | NA | NA |
| BLSA | 1114 | 65.44±14.11 | 589/53 | 729 | 9 | 11 | NA | NA | NA | NA | 365 |
| CARDIA | 892 | 51.21±3.98 | 471/53 | 620 | NA | NA | NA | NA | NA | NA | 272 |
| OASIS | 983 | 69.92±9.75 | 557/57 | 759 | 220 | NA | NA | NA | NA | NA | 4 |
| PENN | 807 | 72.63±<br>10.65 | 333/59 | 173 | 294 | 283 | NA | NA | NA | NA | 57 |
| WHIMS | 995 | 69.61±<br>3.64 | 995/100 | 986 | NA | NA | NA | NA | NA | NA | 6 |
| WRAP | 116 | 63.36±<br>6.06 | 79/68 | 116 | NA | NA | NA | NA | NA | NA | NA |
| PHENOM | 2125 | 30.21±<br>10.60 | 854/40 | 1031 | NA | NA | 1094 | NA | NA | NA | NA |
| ABIDE | 1220 | 17.92±<br>9.01 | 203/17 | 623 | NA | NA | NA | 597 | NA | NA | NA |
| **Replication**<br>**set<sup>a</sup>** | 18,259 | 54.70±<br>7.43 | 9742/53 | NA | NA | NA | NA | NA | NA | NA | NA |

- ADNI: 63 sites (USA)
- UKBB: 5 sites (UK)
- AIBL: 2 sites (Australia)
- BIOCARD: 2 sites (USA)
- BLSA: 1 site (USA)
- CARDIA: 3 sites (USA)
- OASIS: 1 site (USA)
- PENN: 1 site (USA)
- WHIMS: 14 sites (USA)
- WRAP 1 site (USA)
- PHENOM: 12 sites (China, Brazil, Australia, Germany, Spain, USA, Netherlands)
- ABIDE: 25 sites (USA, Netherlands, Belgium, Germany, Ireland, Switzerland, France) Abbreviations: CN: healthy control; AD: Alzheimer’s disease; MCI: mild cognitive impairment; SCZ: schizophrenia; ASD: autism spectrum disorder; MDD: major depressive disorder; DM: diabetes; HTN: hypertension. ^a^UKBB data were separately downloaded two times: the first was the *N*=21,305 in the discovery set, and the second was the replication set. ^b^We define CN (healthy controls) as participants that do not have any of the diseases listed here. These CN participants might have diagnoses of other illnesses or comorbidities (e.g., participants from UKBB have a wide range of pathology based on ICD-10).

**eTable 2:** Clinical phenotypes and diagnoses used in machine learning classification. . We harmonized the population of the phenotypes of interest per study definitions:

| Trait (ICD-10 code or ID) | Sample size (CN/PT or *N*) | Site | Trait (ICD-10 code or ID) | Sample size (CN/PT or *N*) | Site |
| --- | --- | --- | --- | --- | --- |
| AD | 1095/723 | ADNI, PENN, & AIBL | Carpal tunnel syndrome (G560) | 901/901 | UKBB |
| MCI | 1273/1095 | ADNI, PENN, & AIBL | Lesion of ulnar nerve (G562) | 104/104 | UKBB |
| SCZ | 1031/1094 | PHENOM | Lesion of plantar nerve (G576) | 163/163 | UKBB |
| DM | 1093/1093 | UKBB | Angina pectoris (I20) | 1535/1535 | UKBB |
| MDD<sup>a</sup> | 1476/1476 | UKBB | Acute myocardial infarction (I21) | 769/769 | UKBB |
| HTN | 934/887 | ADNI, BLSA & CARDIA | Chronic ischaemic heart disease (I25) | 2217/2217 | UKBB |
| ASD | 623/597 | ABIDE | Pulmonary embolism (I20) | 351/351 | UKBB |
| Iron deficiency anemia (D50) | 1012/1012 | UKBB | Cardiomyopathy (I42) | 116/116 | UKBB |
| Vitamin B12 deficiency anemia (D50) | 78/78 | UKBB | Paroxysmal tachycardia (I47) | 320/320 | UKBB |
| Agranulocytosis (D70) | 245/245 | UKBB | Heart failure (I50) | 436/436 | UKBB |
| Thyrotoxicosis (E05) | 205/205 | UKBB | Cerebral infarction (I63) | 291/291 | UKBB |
| Vitamin D deficiency (E55) | 180/180 | UKBB | Vitamin B deficiency (E53) | 130/130 | UKBB |
| Obesity (E66) | 1481/1481 | UKBB | Hemiplegia (G81) | 111/111 | UKBB |
| Lipoprotein metabolism disorder (E78) | 3880/3880 | UKBB | Facial nerve disorders (G51) | 95/95 | UKBB |
| Mineral metabolism disorder (E83) | 291/291 | UKBB | Tower rearranging (21004) | 8412 | UKBB |
| Volume depletion | 240/240 | UKBB | Matrix pattern (6373) | 8501 | UKBB |
| Delirium | 92/92 | UKBB | TMT-A (6348) | 8599 | UKBB |
| Alcohol abuse | 341/341 | UKBB | TMT-B (6350) | 8599 | UKBB |
| Tobacco abuse | 863/863 | UKBB | DSST (23324) | 8523 | UKBB |
| Bipolar affective disorder | 77/77 | UKBB | Pairs matching (399) | 20945 | UKBB |
| Phobic anxiety disorder | 84/84 | UKBB | Numerical memory (4282) | 9323 | UKBB |
| Multiple sclerosis | 109/109 | UKBB | Prospective memory (4288) | 19681 | UKBB |
| Epilepsy | 250/250 | UKBB | Reaction time (20023) | 21258 | UKBB |
| Migraine | 508/508 | UKBB | Fluid intelligence (20016) | 19184 | UKBB |
| Sleep disorders | 590/590 | UKBB |  |  |  |

- We combined AD and MCI patients from ADNI, PENN, and AIBL but excluded OASIS subjects because of the different diagnostic criteria of an AD patient in OASIS.
- For several binary disease phenotypes, we used the ICD-10 diagnosis (https://biobank.ndph.ox.ac.uk/ukb/field.cgi?id=41270). Note that ICD-10 diagnoses are generally collected from the participants’ medical inpatient records. We first included diseases from the following categories:

◦ Diseases of the blood and blood-forming organs and certain disorders involving the immune mechanism (D-XXX, XXX represents the ID of a specific disease);
◦ Endocrine, nutritional, and metabolic diseases (E-XXX);
◦ Mental and behavioral disorders (F-XXX);
◦ Diseases of the nervous system (G-XXX);
◦ Diseases of the circulatory system (I-XXX).
- then set a threshold of 75 patients for any ICD-10 diagnosis. We finally randomly selected age and sex-matched healthy controls (excluding all patients in all diagnoses). ^a^: For major depressive disorder, we used the inclusion criteria from our previous work.^75^
- For cognitive scores, we included:

◦ Tower rearranging (https://biobank.ndph.ox.ac.uk/showcase/field.cgi?id=21004)
◦ Matrix pattern (https://biobank.ndph.ox.ac.uk/showcase/field.cgi?id=6373)
◦ TMT-A (https://biobank.ndph.ox.ac.uk/showcase/field.cgi?id=6348)
◦ TMT-B (https://biobank.ndph.ox.ac.uk/showcase/field.cgi?id=6350)
◦ DSST (https://biobank.ndph.ox.ac.uk/showcase/field.cgi?id=23324)
◦ Pairs matching (https://biobank.ndph.ox.ac.uk/showcase/field.cgi?id=399)
◦ Numerical memory (https://biobank.ndph.ox.ac.uk/showcase/field.cgi?id=4282)
◦ Prospective memory (https://biobank.ndph.ox.ac.uk/showcase/field.cgi?id=4288)
◦ Reaction time (https://biobank.ndph.ox.ac.uk/showcase/field.cgi?id=20023)
◦ Fluid intelligence (https://biobank.ndph.ox.ac.uk/showcase/field.cgi?id=20016)
- AD: Alzheimer’s disease; MCI: mild cognitive impairment; SCZ: schizophrenia; DM: diabetes mellitus; MDD: major depressive disorder; HTN: hypertension; ASD: autism spectrum disorder; CN: healthy control; PT: patient; *N*: number of participants. We decided not to harmonize cognitive scores from different studies.

**eTable 3:** Comparison of variants identified via MuSIC with other studies. . Using the AAL atlas, we found (using the same data in the current study) that 269 independent significant SNPs had 356 pairwise associations with 54 AAL brain regions. 230 out of the 269 SNPs matched with the SNPs in MuSIC. Among the 39 unmatched SNPs, 15 SNPs were in linkage disequilibrium (LD, *r^2^* > 0.6) with MuSIC SNPs (**Supplementary eFile 5**). As a second example, Zhao et al.^4^ reported that 251 independent significant SNPs had 346 pairwise associations with 43 GM regions using the Mindboggle atlas on the UKBB (*N*=19,629).^82^ 129 of the 251 SNPs matched with SNPs identified by MuSIC. Among these non-matching SNPs (127), 31 were in LD with MuSIC SNPs (**Supplementary eFile 6**). Similarly, Elliot et al.^5^ (*N*=8428) discovered that 20 independent significant SNPs had 58 pairwise associations with 52 GM regions from atlases in Freesurfer and FSL software. Out of the 20 SNPs, 16 coincided with MuSIC SNPs. Among the four unmatched SNPs, 1 SNP was in LD with MuSIC SNPs (**Supplementary eFile 7**). Note that the definition of independent significant SNPs or genomic loci might slightly differ between studies.

| Study/Atlas | Identified genomic loci | Matched loci | Loci in LD | Novel loci | Database | Sample size | Ancestry |
| --- | --- | --- | --- | --- | --- | --- | --- |
| MuSIC | 915 | NA | NA | NA | UKBB | 18,052 | European |
| AAL | 218 | 162 | 13 | 740 | UKBB | 18,052 | European |
| Zhao et al.<sup>4</sup> | 251 | 73 | 14 | 828 | UKBB | 19,629 | European |
| Elliot et al.<sup>5</sup> | 20 | 16 | 1 | 898 | UKBB | 8428 | European |
| GWAS Catalog | NA | 298 | NA | 617 | NA | NA | NA |

**eTable 4:** Classification balanced accuracy for disease classification and effect size of these imaging signatures. Disease classification performance is presented using balanced accuracy. The mean and standard deviation are presented. Cohen’s *d* was computed to compare the SPARE scores between groups. Multi-scale classification^a^: All 2003 PSCs from multiple scales were fit into the classifier. Multi-scale classification^b^: PSCs from all scales were fit into the classifier with a nested feature selection procedure (SVM-REF). The motivation is that PSCs from different scales are hierarchical and correlated. The nested feature selection can select the features most relevant to the specific task. We avoided any statistical comparison of the performance of machine learning models because available statistical tests are liberal and often lead to false-positive conclusions due to the complexity of the cross-validation procedure.^83^

| PSC | AD | $d$ | MCI | $d$ | SCZ | $d$ | DM | $d$ | HTN | $d$ | MDD | $d$ | ASD | $d$ |
| --- | --- | --- | --- | --- | --- | --- | --- | --- | --- | --- | --- | --- | --- | --- |
| C32 | 0.78±0.02 | 1.52 | 0.62±0.02 | 0.59 | 0.55±0.02 | 0.30 | 0.56±0.02 | 0.35 | 0.55±0.02 | 0.28 | 0.52±0.02 | 0.16 | 0.50±0.02 | 0.07 |
| C64 | 0.81±0.02 | 1.73 | 0.63±0.02 | 0.66 | 0.57±0.02 | 0.41 | 0.57±0.02 | 0.40 | 0.56±0.02 | 0.31 | 0.53±0.02 | 0.17 | 0.53±0.02 | 0.19 |
| C128 | 0.82±0.02 | 1.82 | 0.65±0.02 | 0.76 | 0.59±0.02 | 0.47 | 0.56±0.02 | 0.33 | 0.55±0.02 | 0.30 | 0.52±0.02 | 0.15 | 0.52±0.02 | 0.15 |
| C256 | 0.85±0.02 | 2.08 | 0.66±0.02 | 0.91 | 0.59±0.02 | 0.50 | 0.56±0.02 | 0.47 | 0.54±0.02 | 0.31 | 0.51±0.02 | 0.13 | 0.52±0.02 | 0.16 |
| C512 | 0.88±0.02 | 2.34 | 0.67±0.02 | 1.06 | 0.62±0.02 | 0.62 | 0.57±0.02 | 0.54 | 0.56±0.02 | 0.42 | 0.52±0.02 | 0.05 | 0.54±0.02 | 0.24 |
| C1024 | 0.90±0.02 | 2.50 | 0.72±0.02 | 1.12 | 0.65±0.02 | 0.75 | 0.60±0.02 | 0.59 | 0.59±0.02 | 0.46 | 0.56±0.02 | 0.13 | 0.55±0.02 | 0.29 |
| Multi-scale<sup>a</sup> | 0.91±0.02 | 2.54 | 0.72±0.02 | 1.12 | 0.66±0.02 | 0.77 | 0.61±0.02 | 0.64 | 0.59±0.02 | 0.47 | 0.55±0.02 | 0.23 | 0.56±0.02 | 0.30 |
| Multi-scale<sup>b</sup> | 0.92±0.02 | 2.61 | 0.73±0.02 | 1.13 | 0.67±0.02 | 0.78 | 0.64±0.02 | 0.67 | 0.61±0.02 | 0.49 | 0.55±0.02 | 0.26 | 0.58±0.02 | 0.32 |
| AAL | 0.82±0.02 | 1.81 | 0.66±0.02 | 0.75 | 0.59±0.02 | 0.46 | 0.57±0.02 | 0.32 | 0.57±0.02 | 0.35 | 0.52±0.02 | 0.08 | 0.52±0.02 | 0.14 |
| RAVENS | 0.85±0.02 | 2.04 | 0.64±0.02 | 0.74 | 0.60±0.02 | 0.45 | 0.58±0.02 | 0.33 | 0.55±0.02 | 0.34 | 0.50±0.02 | 0.05 | 0.54±0.02 | 0.15 |

| Left-out site | Test BA in CV | Test BA in the left-out site |
| --- | --- | --- |
| ADNI | 0.90±0.02 | 0.88±0.02 |
| AIBL | 0.88±0.02 | 0.95±0.02 |
| PENN | 0.90±0.02 | 0.95±0.02 |

**eTable 5:** 119 MUSE gray matter regions of interest. L: Left hemisphere; R: Right hemisphere; ROI: region of interest.

| MUSE ROI | MUSE ROI | MUSE ROI |
| --- | --- | --- |
| Precentral gyrus (R) | Occipital fusiform gyrus (R) | Anterior insula (L) |
| Precentral gyrus (L) | Planum temporale (R) | Anterior orbital gyrus (R) |
| Accumbens area (R) | Cerebellar vermal lobules I-V | Anterior orbital gyrus (L) |
| Accumbens area (L) | Cerebellar vermal lobules VI-VII | Angular gyrus (R) |
| Amygdala (R) | Cerebellar vermal lobules VIII-X | Angular gyrus (L) |
| Amygdala (L) | Basal forebrain (R) | Calcarine cortex (R) |
| Occipital pole (L) | Basal forebrain (L) | Calcarine cortex (L) |
| Caudate (R) | Middle temporal gyrus (L) | Central operculum (R) |
| Caudate (L) | Occipital pole (R) | Central operculum (L) |
| Cerebellum exterior (R) | Planum temporale (L) | Cuneus (R) |
| Cerebellum exterior (L) | Parietal operculum (L) | Cuneus (L) |
| Planum polare (L) | Postcentral gyrus (R) | Entorhinal area (R) |
| Middle temporal gyrus (R) | Postcentral gyrus (L) | Entorhinal area (L) |
| Hippocampus (R) | Posterior orbital gyrus (R) | Frontal operculum (R) |
| Hippocampus (L) | Temporal pole (R) | Frontal operculum (L) |
| Precentral gyrus medial segment (R) | Temporal pole (L) | Frontal pole (R) |
| Precentral gyrus medial segment (L) | Triangular part of the inferior frontal gyrus (R) | Frontal pole (L) |
| Superior frontal gyrus medial segment (R) | Triangular part of the inferior frontal gyrus (L) | Fusiform gyrus (R) |
| Superior frontal gyrus medial segment (L) | Transverse temporal gyrus (R) | Fusiform gyrus (L) |
| Pallidum (R) | Superior frontal gyrus medial segment (L) | Gyrus rectus (R) |
| Pallidum (L) | Planum polare (R) | Gyrus rectus (L) |
| Putamen (R) | Transverse temporal gyrus (L) | Inferior occipital gyrus (R) |
| Putamen (L) | Anterior cingulate gyrus (R) | Inferior occipital gyrus (L) |
| Thalamus proper (R) | Anterior cingulate gyrus (L) | Inferior temporal gyrus (R) |
| Thalamus proper (L) | Anterior insula (R) | Inferior temporal gyrus (L) |
| Lingual gyrus (R) | Occipital fusiform gyrus (L) | Subcallosal area (R) |
| Lingual gyrus (L) | Opercular part of inferior frontal gyrus (R) | Subcallosal area (L) |
| Lateral orbital gyrus (R) | Opercular part of inferior frontal gyrus (L) | Superior frontal gyrus (R) |
| Lateral orbital gyrus (L) | Orbital part of inferior frontal gyrus (R) | Superior frontal gyrus (L) |
| Middle cingulate gyrus (R) | Orbital part of inferior frontal gyrus (L) | Supplementary motor cortex (R) |
| Middle cingulate gyrus (L) | Posterior cingulate gyrus (R) | Supplementary motor cortex (L) |
| Medial frontal cortex (R) | Posterior cingulate gyrus (L) | Supramarginal gyrus (R) |
| Medial frontal cortex (L) | Precuneus (R) | Supramarginal gyrus (L) |
| Middle frontal gyrus (R) | Precuneus (L) | Superior occipital gyrus (R) |
| Middle frontal gyrus (L) | Parahippocampal gyrus (R) | Superior occipital gyrus (L) |
| Middle occipital gyrus (R) | Parahippocampal gyrus (L) | Superior parietal lobule (R) |
| Middle occipital gyrus (L) | Posterior insula (R) | Superior parietal lobule (L) |
| Medial orbital gyrus (R) | Posterior insula (L) | Superior temporal gyrus (R) |
| Medial orbital gyrus (L) | Parietal operculum (R) | Superior temporal gyrus (L) |
| Superior frontal gyrus medial segment (R) | Posterior orbital gyrus (L) |  |

## eAlgorithm 1 Algorithm for sopNMF.

The source code of the Python implementation of sopNMF is available here: https://github.com/anbai106/SOPNMF

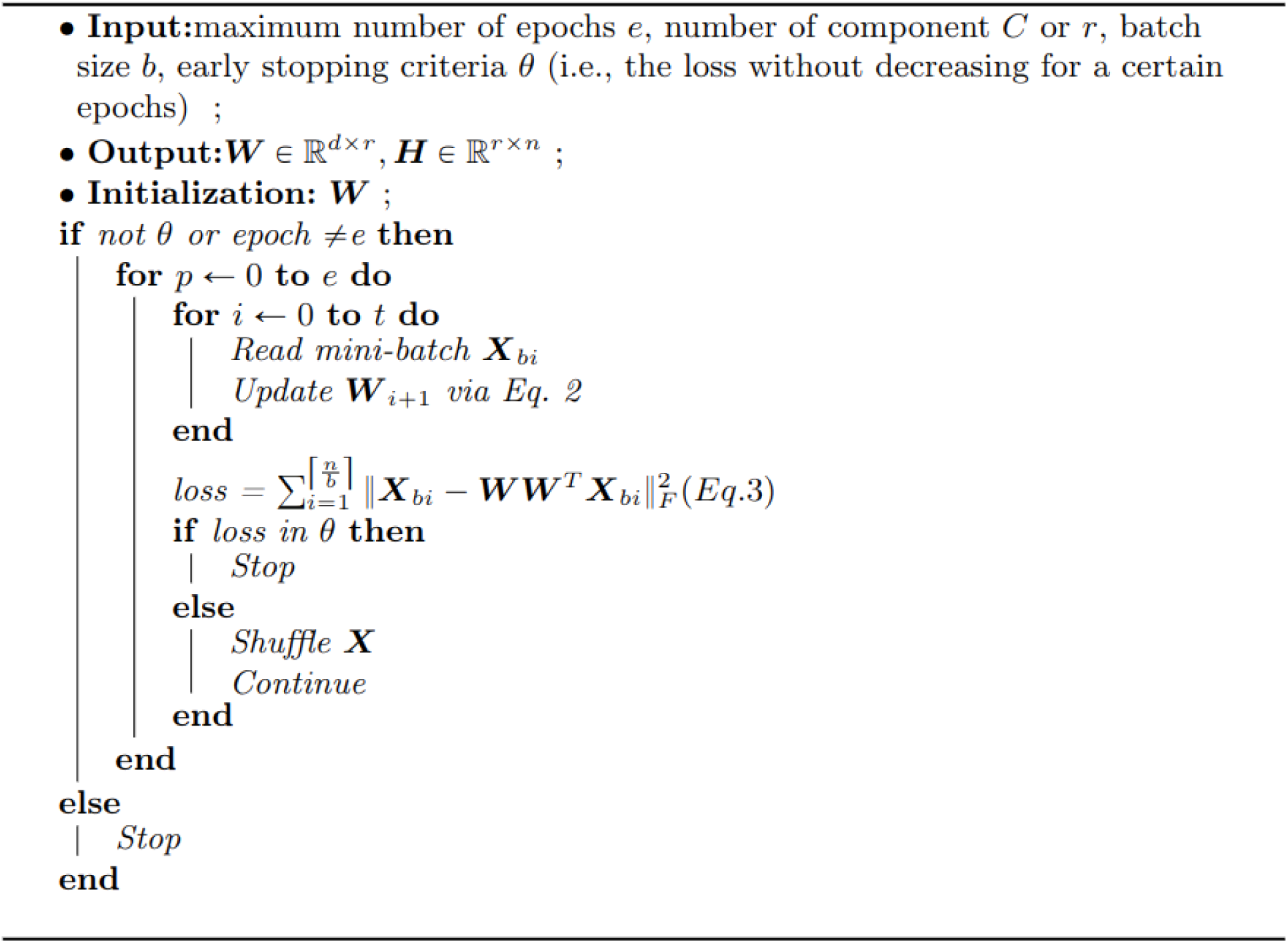

